# Histology-informed microstructural diffusion simulations for MRI cancer characterisation — the Histo-μSim framework

**DOI:** 10.1101/2024.07.15.24310280

**Authors:** Athanasios Grigoriou, Carlos Macarro, Marco Palombo, Daniel Navarro-Garcia, Anna Voronova, Kinga Bernatowicz, Ignasi Barba, Alba Escriche, Emanuela Greco, María Abad, Sara Simonetti, Garazi Serna, Richard Mast, Xavier Merino, Núria Roson, Manuel Escobar, Maria Vieito, Paolo Nuciforo, Rodrigo Toledo, Elena Garralda, Roser Sala-Llonch, Els Fieremans, Dmitry S. Novikov, Raquel Perez-Lopez, Francesco Grussu

## Abstract

Diffusion Magnetic Resonance Imaging (dMRI) simulations in geometries mimicking the complexity of human tissues at the microscopic scale enable the development of innovative biomarkers with unprecedented fidelity to histology. To date, approaches of this kind have focussed heavily on brain imaging. Nevertheless, simulation-informed dMRI has huge potential also in other applications, as for example in body cancer imaging, where new non-invasive biomarkers are still sought. This article fills this gap by introducing a Monte Carlo diffusion simulation framework informed by histology, for enhanced body dMR microstructural imaging — the *Histo-μSim* approach. We generate dictionaries of synthetic dMRI signals with coupled tissue properties from virtual cancer environments, reconstructed from hematoxylin-eosin stains of human liver biopsies. These enable the data-driven estimation of innovative microstructural tissue properties, such as the intrinsic extra-cellular diffusivity, or cell size (CS) distribution moments. We compare *Histo-μSim* to metrics from well-established analytical multi-compartment models *in silico*, on fixed mouse tissues scanned *ex vivo* (kidneys, spleens, and breast tumours) and in cancer patients *in vivo*. Results suggest that *Histo-μSim* is feasible in clinical settings, and that it delivers metrics that more accurately reflect the underlying histology as compared to analytical models. In conclusion, *Histo-μSim* offers histologically-meaningful tissue descriptors that may increase the specificity of dMRI towards cancer, and thus play a crucial role in precision oncology.

## 1 Introduction

The ultimate aim of diffusion MRI (dMRI) is the estimation of statistics of the cellular environment — referred to as *tissue microstructure* — from sets of diffusion-weighted (DW) signal measurements, by solving an inverse mathematical problem [Novikov et al., 2019, Kiselev, 2017]. Multi-compartment biophysical dMRI models have gained momentum as practical approaches capable of providing maps of biologically-meaningful properties, such as cell size (CS) indices. These have found applications in multiple organs, e.g., brain [Veraart et al., 2020], muscles [Fieremans et al., 2017], breast [Xu et al., 2020], liver [Jiang et al., 2020b], prostate [Panagiotaki et al., 2015, Lemberskiy et al., 2018] and beyond. Non-invasive CS measurement may be particularly relevant for disease characterisation and treatment response assessment in oncology, given the variety of cell types that can coexist within tumours, each featuring unique, distinctive dimensions (e.g., normal vs malignant cells, immune cell infiltration, etc) [Reynaud, 2017, Jiang et al., 2020a, Hoffmann et al., 2023, Palombo et al., 2023].

However, current biophysical models are often based on idealised representations of tissue components, such as spheres of fixed radii to describe cells [Panagiotaki et al., 2015, Jiang et al., 2020b, Hoffmann et al., 2023]. This implies that they may neglect other, relevant features of intra-voxel microstructure, e.g., the existence of distributions of CSs, intra-cellular (IC) kurtosis [Lee et al., 2020a, Grussu et al., 2022], or extra-cellular (EC) diffusion time dependence [Burcaw et al., 2015, Xu et al., 2023]. Neglecting such characteristics may bias parameter estimation, and may also cause clinically-relevant information to be missed.

Recently, numerical methods based on more realistic tissue representations have enabled the development of accurate dMRI signal models [Nedjati-Gilani et al., 2017, Lee et al., 2021], increasing the biological specificity of parameter estimation [[Nilsson et al., 2010, Palombo et al., 2016, Palombo et al., 2019, Buizza et al., 2021, Morelli et al., 2023]. In particular, Monte Carlo (MC) diffusion simulations within 3D meshes derived from histology have enabled the characterisation of fine, sub-cellular microstructural details, such as axonal beading/undulation [[Lee et al., 2020b, Lee et al., 2024], or neural process complexity [[Palombo et al., 2016, Palombo et al., 2019]. Nevertheless, to date histology-informed dMRI has focussed heavily on neural tissue, with only a few examples outside the central nervous system [Berry et al., 2018]. More accurate biophysical models are urgently needed in a variety of other contexts beyond brain dMRI, as in oncological body imaging of solid tumours [O’connor et al., 2017]. New dMRI approaches could tackle several, unmet clinical needs, such as patient stratification for treatment selection, response assessment in immunotherapy [Pilard et al., 2021], or the determination of the malignity of lesions that cannot be biopsied [Doblas et al., 2013].

This article aims to fill this gap by introducing a histology-informed MC framework for microstructural diffusion simulations and parameter mapping, referred to as *Histo-μSim*. We present a rich database of virtual cellular environments reconstructed from hematoxylin and eosin (HE) stains of liver biopsies, and use these to synthesise signals for clinically feasible dMRI protocols. The database provides the community with reference values of key cellular properties in cancerous and non-cancerous tissues — information not easily accessible in the literature, yet essential to inform the development of the new dMRI techniques of tomorrow. The set of cellular-level characteristics and corresponding dMRI signals allowed us to devise a strategy for the numerical estimation of unexplored tissue properties with clinically feasible acquisitions. In particular, we tested the estimation of the intrinsic EC diffusivity and of CS distribution moments, which we showcase in pre-clinical scans of fixed mouse tissues and in cancer patients *in vivo*. Results from in *in silico, ex vivo* and *in vivo* data suggest that *Histo-μSim* enables the computation of microstructural metrics that more accurately reflect the underlying histology than standard analytical signal models, and that these can be obtained in clinically acceptable times. In summary, *Histo-μSim* is as a promising new approach for the non-invasive characterisation of body cancers, and may play a crucial role in both clinical practice and research settings, enhancing precision oncology.

## 2 Results

### 2.1 The virtual tissue environments enable histologically-realistic diffusion simulations

We reconstructed 18 virtual tissue environments from regions-of-interest (ROIs) of HE stains of liver tumour biopsies, which we will refer to as *substrates*. The environments enable the generation of synthetic dMRI signals through a MC diffusion simulations, based entirely on open-source software (Fig. 1). The substrates include tissue from non-cancerous liver parenchyma, as well as from primary and metastatic cancers of the liver, such as: primary hepatocellular carcinoma (HCC); metastatic colorectal cancer (CRC); melanoma; breast cancer. The cancer environments encompass a rich variety of cytoarchitectures, including areas of active tumour with high cell density; areas rich in desmoplastic stroma or fibrosis; areas of necrosis; a mix of all of those, as well as regions at the tumour-liver interface. High-resolutions images of the substrates are shown in Supplementary Fig. S2, S3 and S4. In practice, the virtual tissue environments are represented through triangular meshes derived from the outline of cellular structures identified on HE images. As a first demonstration, the environments effectively consist of 3D structures with cylindrical geometry, obtained by prolonging 2D segmentations along the third dimension. dMRI signals are obtained from a substrate through MC simulations, in which water molecules are seeded uniformly within each substrate in both intra-cellular (IC) and extra-cellular (EC) spaces. Afterwards, molecules experience Brownian random walks, simulating diffusion, during which they interact with the boundaries of the cellular structures through elastic reflection. We simulated 100 realisation of each substrate by varying the intrinsic IC and EC diffusivities (*D*_0|*in*_ and *D*_0|*ex*_), obtaining a total of 1800 microstructures.

**Figure 1.**
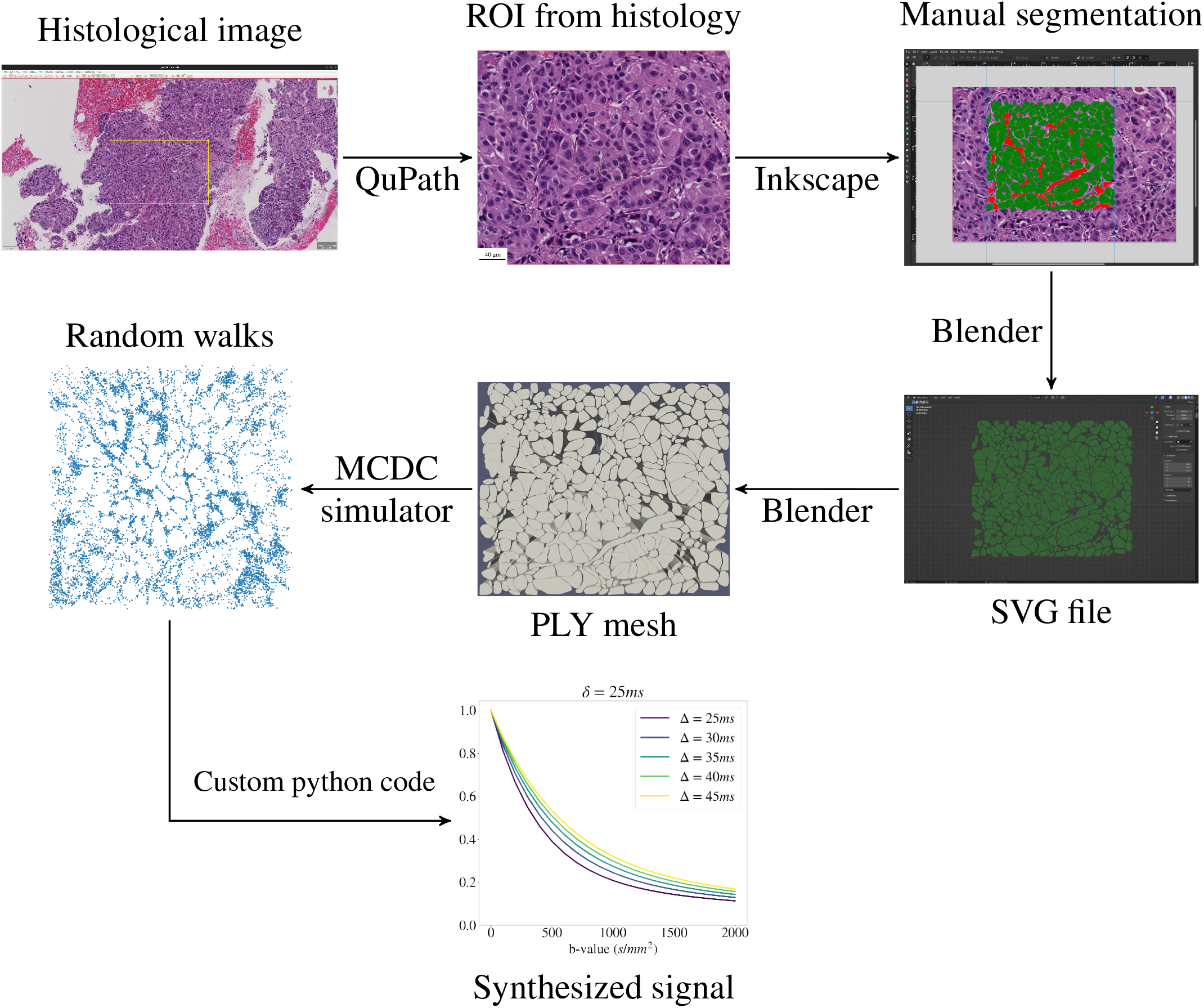
Illustration of our MC simulation framework generating synthetic dMRI signals from histological images. The framework relies on the following open-source software packages: QuPath, Inkscape, Blender, MCDC.

### 2.2 The virtual tissue environments characterise a variety of tissue microstructures

We characterised the reconstructed tissue environments through several metrics, related to the cell size (CS), cell density and to other IC and EC properties. The metrics were:

- the substrate area and cellularity (number of cells per *mm*^2^);
- the IC area fraction *f*_*in*_;
- the fraction of the EC space occupied by luminal structures *f*_*l*_;
- the diameters of the lumina *d*_*lumen*_, when these are present;
- the mean of the cell size (CS) distribution mCS;
- the variance of the CS distribution varCS;
- the skewness of the CS distribution skewCS;
- two volume-weighted mean CS (vCS) indices, vCS_*sph*_ and vCS_*cyl*_ (for a system with spherical vs cylindrical geometry);
- shape and scale parameters of a gamma-distribution [Assaf et al., 2008], which we fitted to the set of cell diameters {*d*_*cell*,1_, *d*_*cell*,2_, …} (see Supplementary Methods for details).

vCS_*sph*_ and vCS_*cyl*_ provide a characteristic CS for the substrate, similarly to mCS. However, as compared to mCS, they put more emphasis on larger cells, and are thus more direct counterparts of dMRI-derived cell size statistics compared to mCS [Veraart et al., 2020, Grussu et al., 2022] (note that in dMRI large cells contribute more to the measured signals than small cells, since they contain more water).

Table 1 reports value of the metrics for all substrates. The reconstructed cancer substrates encompass a rich variety of cytoarchitectures, showing large between-substrate contrasts in all histological metrics. For example, *f*_*in*_ values as high as 0.868 are seen in areas featuring densely packed cells, as in the non-cancerous liver parenchyma, while *f*_*in*_ as low as 0.130 in seen in CRC fibrosis, or as low as 0.024 in necrosis. The table also highlights contrasts in terms of CS. The largest cells are found in the non-cancerous liver (mCS around ~16 *μm*), while all cancers feature the presence of smaller cells. Differences in CS are also seen within the same type of cancer, e.g., mCS of ~13 *μm* and ~6 *μm* in two different CRC substrates. Substrates also feature different skewnesses of the CS distribution, with positive skewCS in most cancers, and negative skewCS in the non-cancerous liver. Finally, in some substrates (e.g., CRC) the EC space features the presence of large lumina, with equivalent diameters as large as ~90 *μm*. Substrates also include areas of partial volume between non-cancerous hepatocytes and cancer cells (substrates 10, 11, 12) with different proportions, a fact that is reflected in different values of skewCS.

**Table 1:**
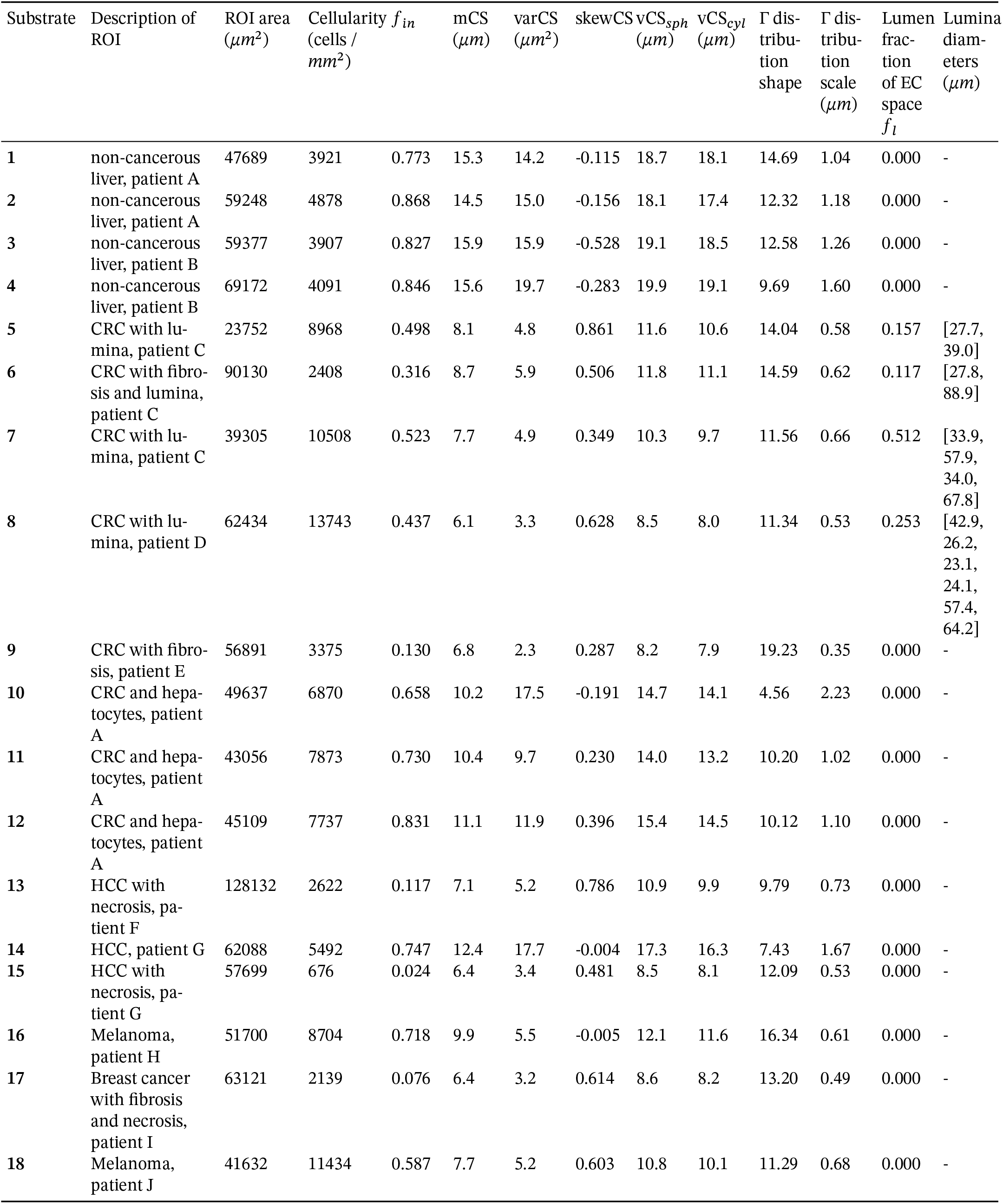
Properties of the substrates used for MC simulations, drawn on histological tissue coming from liver tumour biopsies from 10 cancer patients. Substrates 1 to 4 contain only non-cancerous hepatocytes. Substrates 5 to 9 contain colorectal cancer (CRC) tissue, featuring large luminal spaces or presence of fibrosis, as in substrate 6 and especially substrate 9. Substrates 10 to 12 contain a mix of non-cancerous and different concentrations of CRC cells. Substrates 13 to 15 are sampled from hepatocellular carcinoma (HCC) cases, with substrate 13 in particular containing HCC cells and necrosis, and substrate 15 taken from an area of extended necrosis with scattered cells and cell debris. Substrates 16 and 18 are ROIs of a melanoma liver metastasis. Finally, substrate 17 was taken from a breast cancer liver metastasis, and shows areas of necrosis and fibrosis. The patients’ ID was randomly generated.

Fig. 2 illustrates the different cellular structures that have been identified on HE histology to enable the substrate reconstruction. These are shown in four representative substrates, namely: non-cancerous liver, CRC, breast cancer, and melanoma. The figure highlights again the richness of microstructural characteristics included in our substrates. Tightly packed cells are seen in both non-cancerous liver and in melanoma, with the former showing much larger cells than the latter (mCS of almost 16 *μm* in non-cancerous liver, twice as large as the approximately 8 *μm* seen in melanoma). A wide range of IC fraction *f*_*in*_ is also seen, ranging from 0.076 in the breast cancer substrate (containing fibrotic areas and extensive necrosis) up to 0.846 for the non-cancerous liver. Finally, large luminal spaces in CRC substrates occupy a considerable portion of EC space, with areas equivalent to the space taken by hundreds of cells.

**Figure 2.**
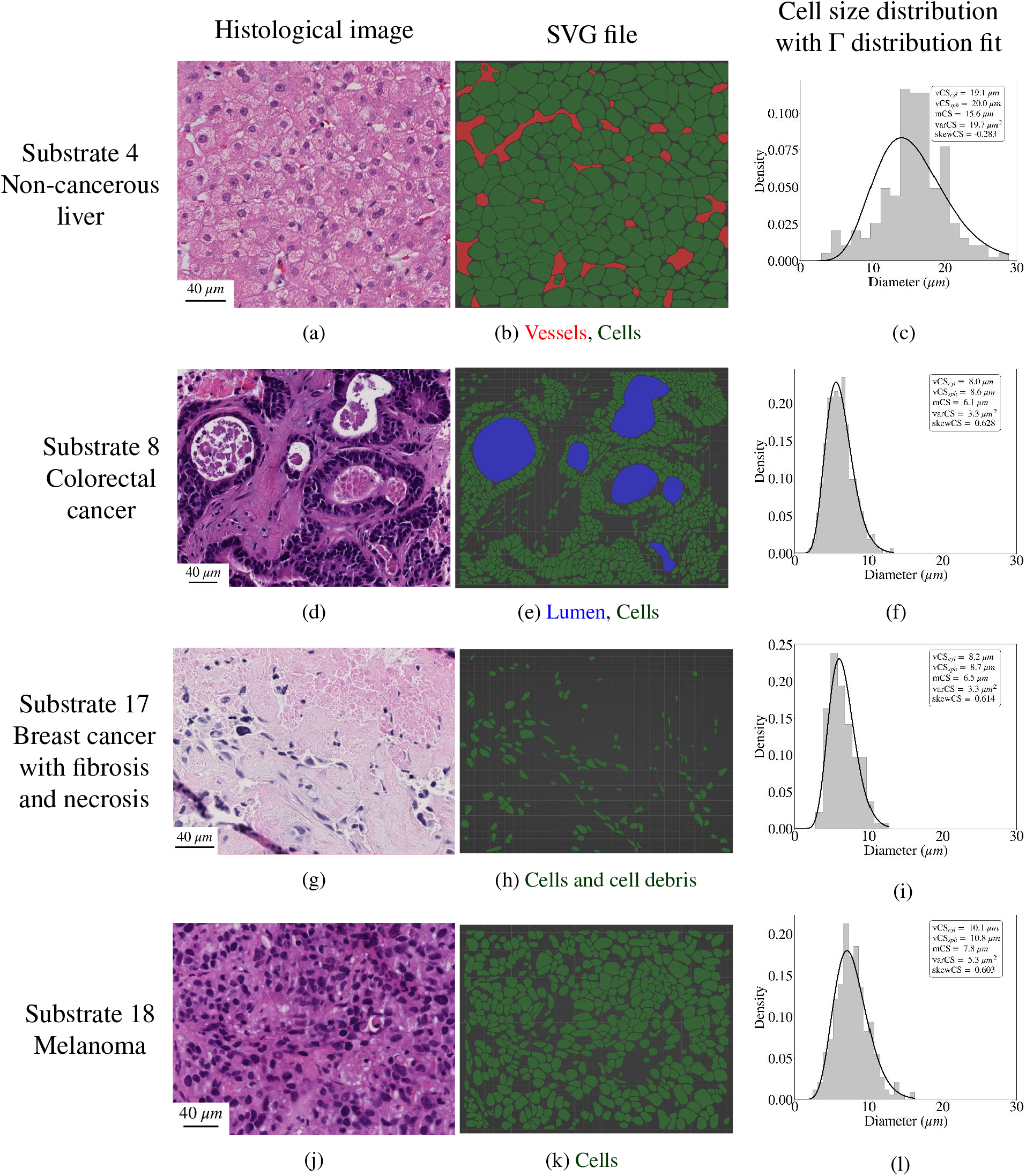
Visualisation of four illustrative substrates used for MC diffusion simulations. Left column (panels (a), (d), (g), (j)): HE histological images. Central columns (panels (b), (e), (h), (k)): SVG files reconstructed with the Blender software package, showing different substrate features (e.g., cells and debris in green, vessels in red, lumina in dark blue). Right column (panels (c), (f), (i), (l)): histograms depicting the CS (i.e., cell diameter) distribution for each substrate, with summary statistics and with a Gamma distribution fit superimposed onto it (black solid line). From top to bottom: non-cancerous liver (substrate 4), colorectal cancer (substrate 8), breast cancer (substrate 17), melanoma (substrate 18).

### 2.3 Histo-μSim parameter estimation outperforms analytical signal modelling

We used the set of paired examples made of synthetic dMRI signals from MC simulations and corresponding histological parameters to inform tissue parameter estimation on unseen dMRI signals. The approach was compared to fitting of a well-established, multi-exponential analytical dMRI signal model, which accounts for restricted IC diffusion within cylindrical structures (given the cylindrical symmetry of our substrates), as well as hindered, EC diffusion [Panagiotaki et al., 2015, Jiang et al., 2020b] (see Methods, Eq. 5). Results unequivocally suggest that our proposed, MC-informed parameter estimation strategy outperforms the fitting of an analytical signal models, since the former provides tissue parameter estimates that correlate more strongly to ground truth values than the latter. For this experiment, we built a MC-informed forward signal model (referred to as *forward model 1*) taking vCS_*cyl*_, *f*_*in*_, *D*_0|*in*_ and *D*_0|*ex*_ as input tissue parameters, being these the same tissue parameters of the analytical signal model.

Fig. 3 shows scatter density plots of ground truth versus estimated tissue parameters in *in silico* experiments. The figure refers to the analysis of dMRI signals synthesised with a pulsed-gradient spin echo (PGSE) protocol matching that of available *in vivo* scans, and referred to as protocol PGSE-in (see Materials and Methods). The protocol include multiple b-values (maximum b = 1500 *s*/*mm*^2^) and multiple diffusion times. It is apparent that *f*_*in*_ and *D*_0|*in*_ are, respectively, the metrics that are the most/the least accurately predicted. Correlation coefficients between ground truth and predicted values are consistently higher for *Histo-μSim* than for analytical modelling. While for both models a strong correlation between estimated and ground truth is seen for *f*_*in*_, a moderate correlation is seen for vCS_*cyl*_ for MC-informed fitting (*r* = 0.57), and a low correlation for the analytical model (*r* = 0.20). For *D*_0|*in*_ instead, it is weak for both approaches, although considerably higher for *Histo-μSim* (*r* = 0.19 against 0.04). Interestingly, we also observe a moderate correlation between ground truth and predicted *D*_0|*ex*_ for MC-informed fitting (*r* = 0.49). Note that the analytical model in Eq. 5 enables the estimation of the EC apparent diffusion coefficient (ADC) ADC_*ex*_, and not of the intrinsic EC diffusivity *D*_0|*ex*_.

**Figure 3.**
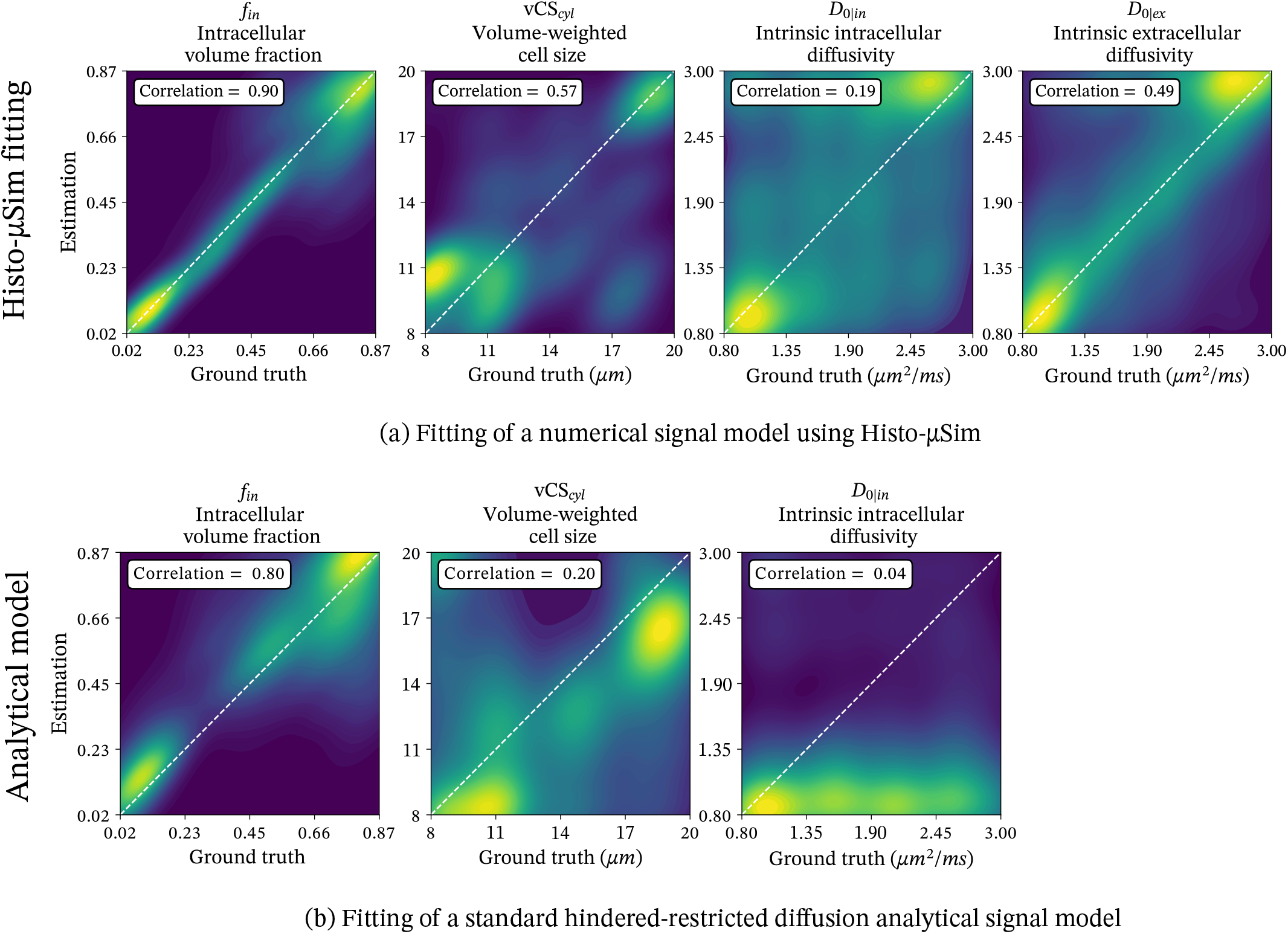
Scatter density plots between ground truth and estimated tissue parameters for MC-informed parameter estimation (*forward model 1*; panel (a) on top) and for standard non-linear least square fitting of a related hindered-restricted diffusion analytical model (panel (b), on the bottom). From left to right: IC fraction *f*_*in*_, CS statistic vCS_*cyl*_, intrinsic IC diffusivity *D*_0|*in*_, intrinsic EC diffusivity *D*_0|*ex*_ (only for MC-informed fitting on top). The plots also include the identity line for reference, and the Pearson’s correlation coefficient between ground truth and estimated parameter values. Results are shown for dMRI protocol PGSE-in.

### 2.4 Histo-μSim enables data-driven CS distribution characterisation

We also investigated whether the *Histo-μSim* framework enables the data-driven, equation-free estimation of additional features of the CS distribution, namely its mean, variance and skewness (mCS, varCS and skewCS). This experiment was motivated by the fact that in our substrates, these CS distribution moments are encoded in the IC signal decay. This is visualised in Fig. 4, in which a clear modulation of the IC ADC ADC_*in*_ is seen as mCS, varCS and skewCS vary. The dependence of ADC_*in*_ on the CS distribution moments is approximately linear for mCS and varCS.

**Figure 4.**
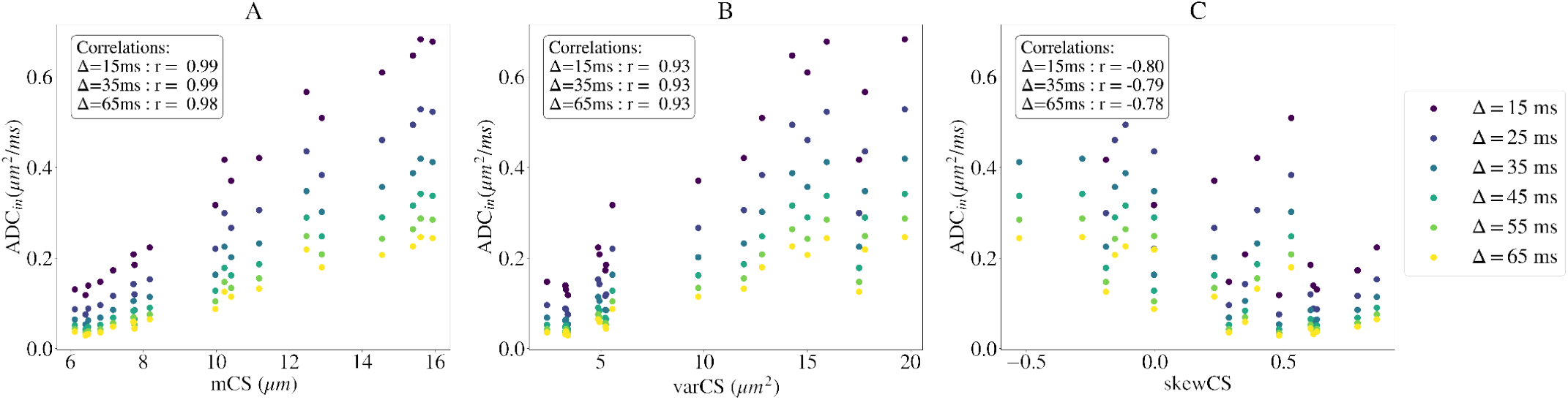
Relation between the IC ADC of an ideal PGSE experiment and CS distribution moments, shown for *D*_0|*in*_ = 2.2 *μm*^2^/*ms* and fixed gradient duration *δ* = 15 *ms*, with increasing gradient separation Δ.

We tested MC-informed fitting of a second signal model, referred to as *forward model 2*, with tissue parameters mCS, varCS, skewCS, *f*_*in*_, *D*_0|*in*_ and *D*_0|*ex*_. Fig. 5 shows parameter estimation results for the same PGSE-in protocol used previously in Fig. 3. Supplementary Fig. S5 and S6 report instead results from simulation-informed fitting of forward model 2 for two additional dMRI protocols, namely: a DW twice-refocussed spin echo (TRSE) acquisition, matching that of another set of available *in vivo* dMRI scans (maximum b-value: 1600 *s*/*mm*^2^); a second PGSE acquisition, matching an high-field acquisition performed on fixed *ex vivo* mouse tissue (maximum b-value: 4500 *s*/*mm*^2^). We will refer to the former as protocol TRSE, while to the latter as protocol PGSE-ex.

**Figure 5.**
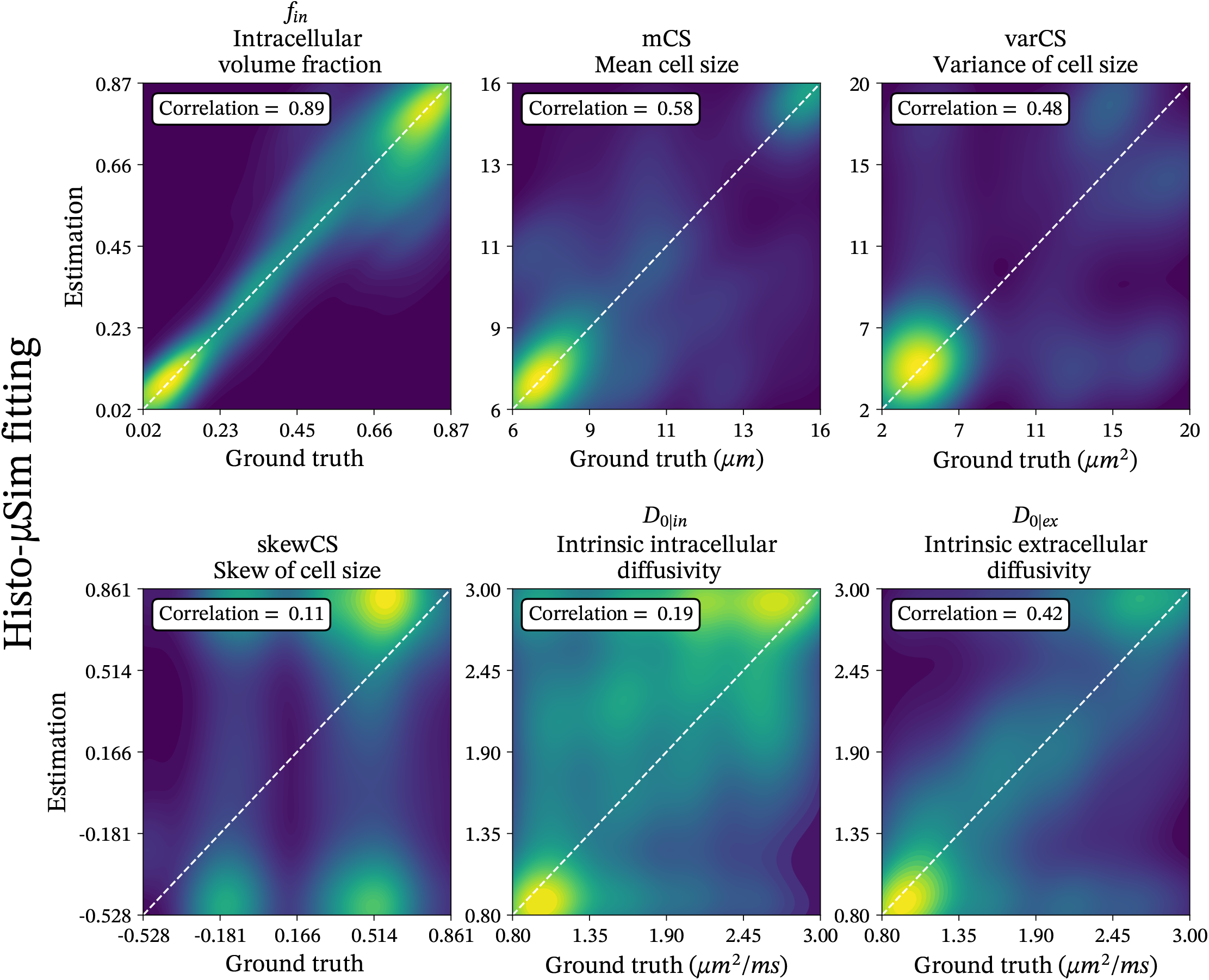
Scatter density plots between ground truth and estimated tissue parameters for MC-informed parameter estimation (forward model 2) and dMRI protocol PGSE-in. Each plot corresponds to a metric. From the top left corner, in clock-wise order: IC fraction *f*_*in*_, mean CS index, variance of CS varCS, intrinsic EC diffusivity *D*_0|*ex*_, intrinsic IC diffusivity *D*_0|*in*_, skewness of CS distribution skewCS. The plots also include the identity line for reference, and the Pearson’s correlation coefficient between ground truth and estimated parameter values. Results are shown for dMRI protocol *PGSE-in*.

Findings from all protocols converge towards the feasibility of estimating mCS and varCS, given that moderate-to-strong correlations are seen between ground truth and estimated parameter values for these metrics. Regarding skewCS, estimated value retain certain sensitivity to the reference ground truth, and appear capable of resolving at least strong contrasts between negative and positive skewCS values. However, for this metrics the quality of the estimation depends strongly on the protocol. In more detail, mCS is the metric that is most accurately predicted, with correlations varying from moderate (*r* = 0.58 for PGSE-in) to strong (*r* = 0.71 for TRSE and 0.83 for PGSE-ex). For varCS we observe moderate correlations between estimated and ground truth values (up to *r* = 0.69 for PGSE-ex), while for skewCS correlations are weaker (highest correlation of *r* = 0.20 for TRSE). Results for the estimation of *f*_*in*_, *D*_0|*in*_ and *D*_0|*ex*_ are in line with what was seen for *forward model 1* (good agreement for *f*_*in*_ in all cases, with highest correlation *r* = 0.94 for TRSE; weaker correlations for *D*_0|*in*_, with highest *r* = 0.30 for PGSE-ex; weak-to-moderate correlations for *D*_0|*ex*_, with highest correlation *r* = 0.42 for PGSE-in).

### 2.5 Histo-μSim microstructural parameters correlate with their histological counterparts in fixed mouse tissue

We tested *Histo-μSim* fitting on pre-clinical PGSE scans, acquired at 9.4T on 8 formalin-fixed *ex vivo* mouse tissue specimens, for which HE sections were also available. These were: a non-cancerous breast and 3 breast tumours from the mouse mammary tumour virus (MMTV) polyomavirus middle T antigen (PyMT) transgenic mouse model [Guy et al., 1992, Attalla et al., 2021], obtained at weeks 9, 11 and 14; a normal spleen and a spleen suffering from splenomegaly from the MMTV mice; two kidneys from C57BL/6 WT male mice (9 weeks old), one normal and one featuring folic acid-induced injury [Yan, 2021]. Quantitative analyses also show that key *Histo-μSim* metrics correlate with their direct histological counterparts, as illustrated by the correlation matrix in Fig. 6. For example, we observe a statistically significant, positive, strong correlation between *Histo-μSim f*_*in*_ and mCS with histological *f*_*in*_ and mCS (*r* = 0.68 between *f*_*in*|*MC*_ and *f*_*in*|*histo*_, *p* = 0.002; *r* = 0.65 between mCS_*MC*_ and mCS_*histo*_, *p* = 0.003), and a moderate, borderline-significant positive correlation between varCS_*MC*_ and varCS_*histo*_ (*r* = 0.43, *p* = 0.077). Conversely, the correlation between *Histo-μSim* and histological skewCS is only weak (*r* = 0.09, *p* = 0.719 between skewCS_*MC*_ and skewCS_*histo*_). These correlations, which are comparable to those obtained previously on *in silico* data (Supplementary Fig. S6), are systematically stronger than those obtained for the analytical signal model, demonstrating the potential of *Histo-μSim* for increasing dMRI biological specificity (Fig. 6: *r* = 0.63, *p* = 0.005 between *f*_*in*|*AN*_ and *f*_*in*|*histo*_; *r* = 0.37, *p* = 0.125 between vCS_*AN*_ and vCS_*sph*|*histo*_).

**Figure 6.**
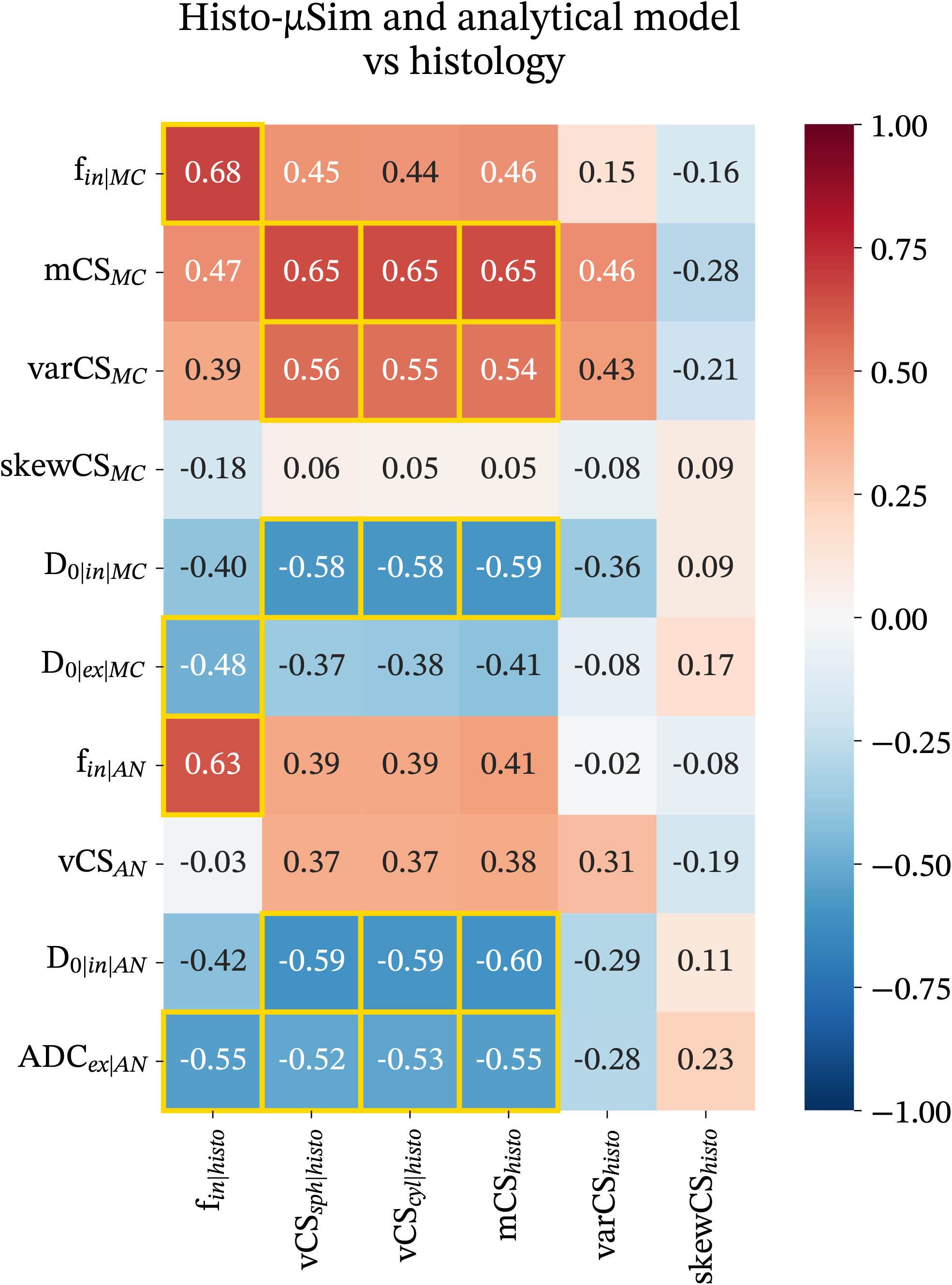
Pearson’s correlation coefficients between all MRI metrics and histological indices obtained on fixed mouse tissue *ex vivo*. Metrics from *Histo-μSim* are indicated by subscript *MC*, for “Monte Carlo simulation-informed”; metrics from the analytical signal models are indicated by subscript *AN*, for “analytical”; metrics from histology are indicated by subscript *histo*. Metrics are: IC fraction *f*_*in*_; mean, variance and skewness of CS (mCS, varCS, skewCS); volume-weighted characteristic CS indices (vCS); intrinsic IC and EC diffusivities (*D*_0|*in*_ and *D*_0|*ex*_); EC ADC (ADC_*ex*_). *p* < 0.05 is flagged by yellow squares. Histological metrics were obtained through manual segmentation of cells on HE data.

Tables 2 and 3 provides dMRI and histological metrics within all ROIs. The values in the tables were used to generate Fig. 6. Contrasts in histological metrics agree with dMRI in several cases. For example, histological mCS is lower in necrotic compared to non-necrotic areas in the week 14 breast tumour (ROI 2 vs 1), or in the normal spleen compared to the normal kidney (ROI 17 compared to ROI 16). Histological *f*_*in*_ is higher in the week 9 breast tumour than in the non-cancerous breast (ROI 1 vs 5), and very low in necrosis (ROI 2). In some cases, differences between dMRI and corresponding histology metrics are also seen, e.g., the low dMRI *f*_*in*_ seen in the healthy kidney underestimates considerably the corresponding *f*_*in*_ values from histology (ROI 16).

**Table 2:** Mean values of metrics from *Histo-μSim forward model 2* and from histology within different ROIs drawn on the breast, kidney and spleen tissue scanned *ex vivo* on a pre-clinical 9.4T MRI system. Histological metrics are indicated with subscript *histo*, while *Histo-μSim* metrics with *MC*, for Monte Carlo simulation-informed estimation.

| Description | $f_{in\vert histo}$ | $f_{in\vert MC}$ | $mCS_{histo}$<br>[$\mu m$] | $mCS_{MC}$<br>[$\mu m$] | $varCS_{histo}$<br>[$\mu m^2$] | $varCS_{MC}$<br>[$\mu m^2$] | $skewCS_{histo}$ | $skewCS_{MC}$ |
| --- | --- | --- | --- | --- | --- | --- | --- | --- |
| ROI 1: Breast tumour – Week 9 (cellular area) | 0.70 | 0.67 | 7.5 | 14.1 | 1.9 | 12.4 | 0.503 | 0.368 |
| ROI 2: Breast tumour – Week 14 (necrosis) | 0.03 | 0.14 | 4.0 | 9.0 | 0.5 | 7.2 | 0.228 | 0.278 |
| ROI 3: Breast tumour – Week 14 (cellular area) | 0.72 | 0.61 | 9.0 | 14.0 | 3.6 | 12.3 | -0.214 | 0.277 |
| ROI 4: Breast tumour - Week 11 (cellular area) | 0.64 | 0.77 | 7.2 | 14.5 | 2.8 | 16.1 | -0.004 | 0.278 |
| ROI 5: Non-cancerous breast | 0.11 | 0.05 | 6.3 | 7.4 | 2.0 | 5.7 | 0.681 | 0.382 |
| ROI 6: Breast tumour – Week 9 (cellular area) | 0.68 | 0.64 | 5.2 | 7.1 | 0.6 | 5.3 | 0.279 | -0.098 |
| ROI 7: Breast tumour – Week 9 (cellular area) | 0.55 | 0.61 | 6.8 | 9.1 | 2.0 | 6.7 | 0.253 | 0.473 |
| ROI 8: Breast tumour – Week 9 (cellular area) | 0.76 | 0.56 | 7.6 | 9.5 | 2.5 | 7.9 | -0.066 | 0.515 |
| ROI 9: Breast tumour - Week 11 (cellular area) | 0.81 | 0.44 | 8.7 | 10.3 | 2.0 | 6.2 | -0.263 | 0.703 |
| ROI 10: Breast tumour - Week 11 (cellular area) | 0.84 | 0.67 | 9.9 | 15.0 | 2.5 | 13.2 | 0.305 | 0.3 |
| ROI 11: Breast tumour - Week 11 (cellular area) | 0.77 | 0.39 | 8.1 | 10.2 | 1.8 | 7.8 | 0.103 | 0.211 |
| ROI 12: Breast tumour - Week 14 (necrosis) | 0.01 | 0.03 | 1.7 | 7.5 | 0.1 | 4.4 | 0.720 | 0.454 |
| ROI 13: Breast tumour - Week 14 (cellular area) | 0.74 | 0.60 | 8.5 | 13.4 | 2.9 | 10.3 | -0.236 | 0.275 |
| ROI 14: Breast tumour - Week 14 (cellular area) | 0.77 | 0.43 | 8.8 | 10.4 | 1.8 | 7.7 | 0.460 | 0.226 |
| ROI 15: Fibrotic kidney | 0.66 | 0.39 | 5.0 | 10.6 | 1.3 | 7.0 | 0.085 | 0.354 |
| ROI 16: Healthy kidney | 0.82 | 0.20 | 10.1 | 10.5 | 6.6 | 8.2 | -0.970 | 0.097 |
| ROI 17: Healthy spleen | 0.75 | 0.36 | 4.4 | 8.0 | 1.0 | 6.9 | 0.244 | 0.051 |
| ROI 18: Splenomegaly | 0.64 | 0.42 | 4.0 | 8.2 | 0.7 | 6.5 | 0.131 | 0.278 |

**Table 3:** Mean values of metrics from the analytical signal model and from histology within different ROIs drawn on the breast, kidney and spleen tissue scanned *ex vivo* on a pre-clinical 9.4T MRI system. Histological metrics are indicated with subscript *histo*, while metrics from the analytical signal model with *AN*, for analytical modelling.

| Description | $f_{in\vert histo}$ | $f_{in\vert AN}$ | $vCS_{sph\vert histo}$<br>[$\mu m$] | $vCS_{cyl\vert histo}$<br>[$\mu m$] | $mCS_{histo}$<br>[$\mu m$] | $vCS_{AN}$<br>[$\mu m$] |
| --- | --- | --- | --- | --- | --- | --- |
| ROI 1: Breast tumour – Week 9 (cellular area) | 0.70 | 0.76 | 8.6 | 8.4 | 7.5 | 16.9 |
| ROI 2: Breast tumour – Week 14 (necrosis) | 0.03 | 0.17 | 4.5 | 4.4 | 4.0 | 15.6 |
| ROI 3: Breast tumour – Week 14 (cellular area) | 0.72 | 0.65 | 10.4 | 10.1 | 9.0 | 16.5 |
| ROI 4: Breast tumour - Week 11 (cellular area) | 0.64 | 0.64 | 8.7 | 8.4 | 7.2 | 14.3 |
| ROI 5: Non-cancerous breast | 0.11 | 0.06 | 7.7 | 7.4 | 6.3 | 13.9 |
| ROI 6: Breast tumour – Week 9 (cellular area) | 0.68 | 0.61 | 5.7 | 5.6 | 5.2 | 8.4 |
| ROI 7: Breast tumour – Week 9 (cellular area) | 0.55 | 0.80 | 8.0 | 7.7 | 6.8 | 13.1 |
| ROI 8: Breast tumour – Week 9 (cellular area) | 0.76 | 0.69 | 8.8 | 8.6 | 7.6 | 13.5 |
| ROI 9: Breast tumour - Week 11 (cellular area) | 0.81 | 0.80 | 9.6 | 9.4 | 8.7 | 17.0 |
| ROI 10: Breast tumour - Week 11 (cellular area) | 0.84 | 0.59 | 11.0 | 10.8 | 9.9 | 15.3 |
| ROI 11: Breast tumour - Week 11 (cellular area) | 0.77 | 0.57 | 9.1 | 8.9 | 8.1 | 15.0 |
| ROI 12: Breast tumour - Week 14 (necrosis) | 0.01 | 0.06 | 2.0 | 1.9 | 1.7 | 15.1 |
| ROI 13: Breast tumour - Week 14 (cellular area) | 0.74 | 0.63 | 9.7 | 9.5 | 8.5 | 15.8 |
| ROI 14: Breast tumour - Week 14 (cellular area) | 0.77 | 0.55 | 9.8 | 9.5 | 8.8 | 14.4 |
| ROI 15: Injured kidney | 0.66 | 0.63 | 6.0 | 5.8 | 5.0 | 17.0 |
| ROI 16: Healthy kidney | 0.82 | 0.05 | 11.8 | 11.6 | 10.1 | 15.0 |
| ROI 17: Healthy spleen | 0.75 | 0.39 | 5.2 | 5.1 | 4.4 | 10.7 |
| ROI 18: Splenomegaly | 0.64 | 0.32 | 4.7 | 4.6 | 4.0 | 11.2 |

Fig. 7 shows examples of dMRI and co-localised HE images in the four breast specimens. These contain a variety of cytoarchitectural environments, with higher inter-sample and intra-sample heterogeneity. For example, the non-cancerous breast features areas rich in stroma. Conversely, higher cell densities are observed in the three MMTV-PyM tumours. At late stages (week 14 tumour), widespread necrosis is also seen. Fig. 8 shows parametric maps from forward model 2 (namely: *f*_*in*_, *D*_0|*in*_, mCS, varCS, skewCS and *D*_0|*ex*_) in the same breast specimens. The variability of cellular microarchitectures seen in Fig. 7 is reflected in the parametric maps. For example, reduced *f*_*in*_ is seen in areas compatible with necrosis within the week 14 tumour (ROI 2, Fig. 8). Additionally, higher *f*_*in*_ is seen in the week 11 tumour, compared to the non-cancerous breast. On histology, this contrast corresponds to presence of areas featuring high cellularity (Fig. 8, ROI 4), compared to stroma in the non-cancerous breast (Fig. 8, ROI 5). Changes in CS moments with respect to the non-cancerous breast are also seen, e.g., reduced mCS and varCS in areas compatible with presence of cell debris in necrosis (ROI 2, Fig. 8). Local variations of IC and EC diffusivities *D*_0|*in*_ and *D*_0|*ex*_ are also seen. For example, *D*_0|*in*_ is lower in areas with high *f*_*in*_ (e.g., in ROI 4 in the week 14 tumour), and *D*_0|*ex*_ is the highest at the interface between specimens and the agarose.

**Figure 7.**
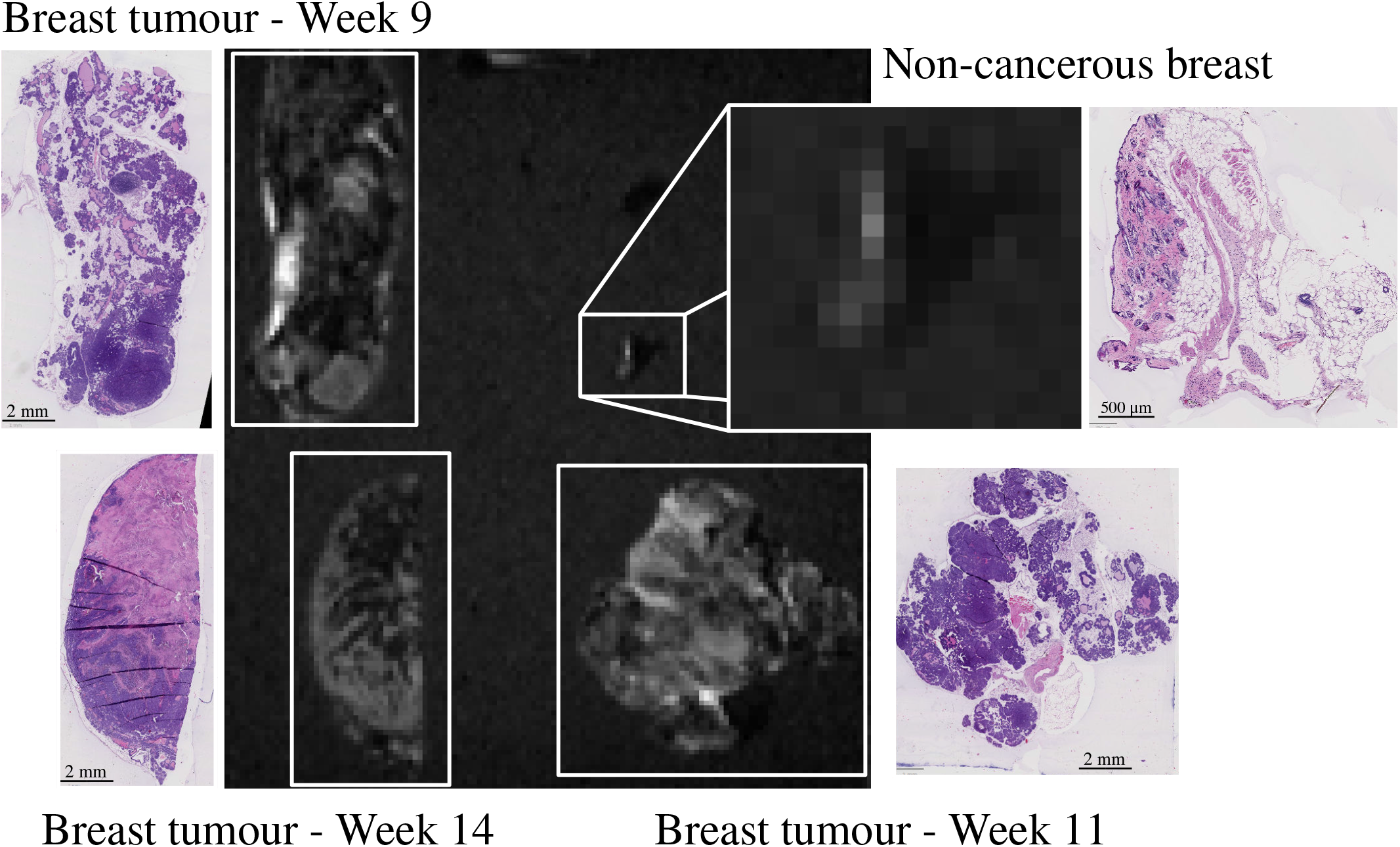
Images from the breast tissue samples that were scanned *ex vivo* at 9.4T. From top left, clock-wise: week 9 MMTV-PyM breast tumour, non-cancerous breast, week 11 MMTV-PyM breast tumour, week 14 MMTV-PyM breast tumour. For each specimen, the b = 0 dMR image and the co-localised HE-stained section are shown.

**Figure 8.**
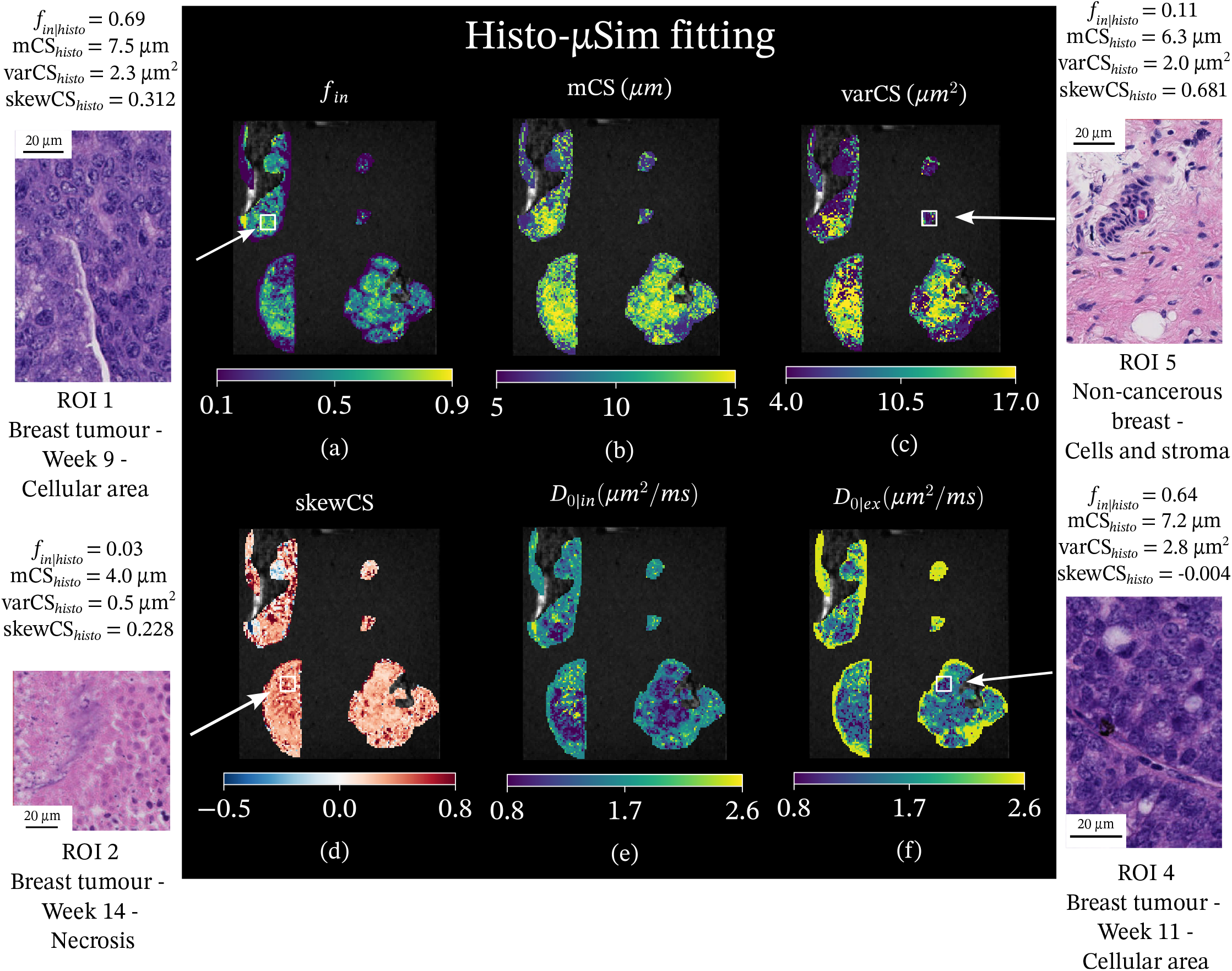
Parametric maps from *Histo-μSim forward model 2* as obtained on the mouse breast specimens scanned *ex vivo* on a 9.4T system. First row: IC fraction *f*_*in*_ (a); mean CS index mCS (b); variance of CS varCS (c). Second row: skewness of the CS distribution skewCS (d); intrinsic IC diffusivity *D*_0|*in*_ (e); intrinsic EC diffusivity *D*_0|*ex*_ (f). For each metric, we show results on the four breast specimens. Moving clock-wise: week 9 MMTV-PyM breast tumour (top left), non-cancerous breast (top right), week 11 MMTV-PyM breast tumour (bottom right), week 14 MMTV-PyM breast tumour (bottom left). Examples of histological tiles in different ROIs are also included, alongside with corresponding quantitative histological indices for each ROIs.

Supplementary Fig. S7 reports maps of microstructural parameters from analytical signal model fitting in the mouse breast specimens. Maps contrasts generally match those from *Histo-μSim* fitting, and highlight similar microstructural characteristics (e.g., necrosis in the week 14 MMTV breast tumour).

Supplementary Fig. S8 shows *Histo-μSim* maps and HE data in a normal spleen and in splenomegaly secondary to late-stage MMTV tumour growth. The spleens exhibit a patchy structure in most dMRI metrics. The same pattern is seen in HE histology, where an alternation of white and red pulps is seen (white pulps known to contain higher T-cell density than red pulps, which are instead rich in blood and iron). Supplementary Fig. S9 shows results from the two kidney samples: one normal, and one following folic-acid induced injury. On histology, the former shows normal representation of all kidney structures, while the injured case shows proximal tubule alteration and extensive inflammation. In terms of dMRI metrics, the injured kidney shows increased *f*_*in*_ and reduced *D*_0|*in*_ and *D*_0|*ex*_ as compared to the normal case. Higher *f*_*in*_ is also seen in the injured kidney cortex as compared to its medulla, a finding that corresponds to higher cell density on visual inspection of histology stains.

### 2.6 Histo-μSim is feasible in cancer patients in vivo

Lastly, we tested *Histo-μSim* for tumour characterisation in cancer patients *in vivo*. In this demonstration, we included scans from 27 patients suffering from advanced solid tumours, primary or metastatic. These were scanned at abdominal or pelvic level, on either a clinical 1.5T or 3T MRI scanner, with a 15-minute dMRI protocol (maximum b-value around 1500 *s*/*mm*^2^). Moreover, we also included HE-stained histological material from a biopsy, which was collected from one of the patient’s tumours, approximately one week after MRI. The analysis of the dMRI scans shows that *Histo-μSim* is feasible *in vivo* within clinically acceptable scan times, and that it provides metrics whose intra-tumour and inter-tumour contrasts are compatible with the cellular environments seen on the biopsies. Furthermore, despite the inherent challenge of comparing dMRI maps obtained over large tumoural areas with histological metrics obtained from a tiny slivers of biopsied tissue, MRI-histology correlations show that *Histo-μSim* IC fraction *f*_*in*|*MC*_ and mean CS mCS_*MC*_ are positively correlated with their histological counterparts from the HE images, albeit weakly (Fig. 9: *r* = 0.37 and *p* = 0.059 between *f*_*in*|*MC*_ and *f*_*in*|*histo*_; *r* = 0.23 and *p* = 0.240 between mCS_*MC*_ and mCS_*histo*_). These correlations are stronger than those of a standard analytical signal model (*r* = 0.25, *p* = 0.200 between *f*_*in*|*AN*_ and *f*_*in*|*histo*_; *r* = 0.015, *p* = 0.940 between vCS_*AN*_ and vCS_*sph*|*histo*_, or *r* = 0.073, *p* = 0.720 between vCS_*AN*_ and mCS_*histo*_). All in all, these findings show that *Histo-μSim* has clinical potential, as it may serve as a useful tool for enhanced non-invasive tumour biology characterisation through dMRI in real-world clinical settings.

**Figure 9.**
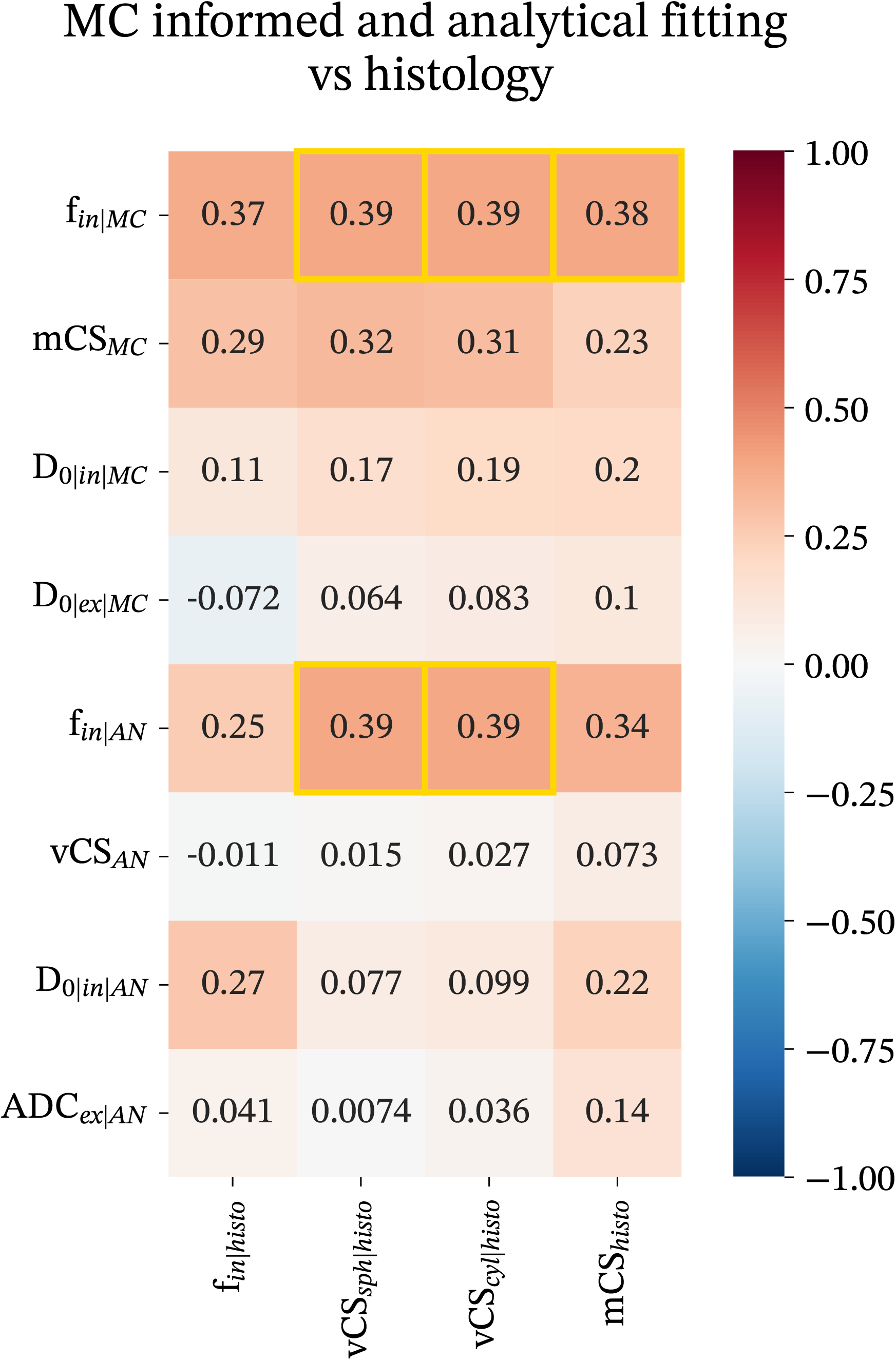
Pearson’s correlation coefficients between all MRI metrics and histological indices obtained on humans *in vivo*. Metrics from *Histo-μSim* are indicated by subscript *MC*, for “Monte Carlo simulation-informed”; metrics from the analytical signal models are indicated by subscript *AN*, for “analytical”; metrics from histology are indicated by subscript *histo*. Metrics are: IC fraction *f*_*in*_; mean CS (mCS) and volume-weighted characteristic CS (vCS) indices; intrinsic IC and EC diffusivities (*D*_0|*in*_ and *D*_0|*ex*_); EC ADC (*ADC*_*ex*_). *p* < 0.05 is flagged by yellow squares. Histological metrics were obtained by automatic image processing in QuPath.

Examples of parametric maps from *Histo-μSim forward model 2* obtained *in vivo* are shown in Fig. 10 in two patients (ovarian cancer liver metastases, scanned at 1.5T; endometrial cancer, scanned at 3T). Maps show intra-tumour variability. For example, in the ovarian cancer case, the largest liver metastasis features reduced *f*_*in*_ and mCS and increased *D*_0|*ex*_ in the necrotic core compared to the tumour outer ring. For the endometrial cancer case, maps reveal different microstructural environments within the tumour, i.e., areas with higher/lower *f*_*in*_, matching areas with lower/higher mCS. Inspection of histological images confirms the existence of heterogeneous cellular characteristics in both cases (Fig. 10), i.e., presence of active cancer and necrosis in the ovarian cancer case, and presence of necrotic areas with abundance of cell debris adjacent to areas with high cellularity in the endometrial tumour. Fitting of a standard analytical model provides metrics that show similar trends, highlight again, for example, the necrotic core in the ovarian cancer metastasis (Supplementary Fig. S10 and S11). Tables and 4 and 5 summarises dMRI and histological metrics within all biopsied tumours. Both dMRI and histology reveal inter-tumour heterogeneity. dMRI-derived values of IC fraction *f*_*in*_ are consistently lower than reference histological *f*_*in*|*histo*_.

**Table 4:** dMRI and histology metrics in cancer patients, with salient patients’ demographic and clinical information. The table reports dMRI metrics from *Histo-μSim*, i.e., fitting of a numerical signal model informed by MC simulations. Patients’ age has been discretised with a granularity of 5-year intervals. HCC stands for hepatocellular carcinoma (primary liver cancer).

| Patient information / MRI scanner | Primary cancer | $f_{in\vert histo}$ | $f_{in\vert MC}$ | $mCS_{histo} [\mu m]$ | $mCS_{MC} [\mu m]$ | $varCS_{MC} [\mu m^2]$ | $skewCS_{MC}$ |
| --- | --- | --- | --- | --- | --- | --- | --- |
| Case 0, 75-80 y.o, F, 3T scanner | colon | 0.58 | 0.24 | 11.4 | 8.9 | 6.3 | 0.288 |
| Case 1, 60-65 y.o, M, 3T scanner | colon | 0.33 | 0.15 | 11.9 | 8.1 | 5.7 | 0.331 |
| Case 2, 60-65 y.o, M, 3T scanner | renal | 0.29 | 0.27 | 9.7 | 8.8 | 6.8 | 0.290 |
| Case 3, 65-70 y.o, M, 3T scanner | colon | 0.23 | 0.17 | 10.9 | 8.3 | 5.9 | 0.287 |
| Case 4, 65-70 y.o, M, 3T scanner | colon | 0.56 | 0.18 | 11.2 | 8.4 | 6.0 | 0.262 |
| Case 5, 70-75 y.o, M, 3T scanner | colon | 0.57 | 0.19 | 11.6 | 8.7 | 5.7 | 0.294 |
| Case 6, 65-70 y.o, M, 3T scanner | colon | 0.47 | 0.20 | 11.5 | 8.5 | 5.6 | 0.337 |
| Case 7, 45-50 y.o, F, 3T scanner | skin cancer | 0.64 | 0.18 | 13.0 | 8.8 | 5.6 | 0.287 |
| Case 8, 75-80 y.o, F, 3T scanner | rectal | 0.57 | 0.10 | 11.4 | 7.5 | 5.0 | 0.147 |
| Case 9, 50-55 y.o, M, 3T scanner | HCC | 0.002 | 0.18 | 6.6 | 8.7 | 5.4 | 0.272 |
| Case 10, 60-65 y.o, F, 3T scanner | ovarian | 0.52 | 0.18 | 13.7 | 8.7 | 6.1 | 0.289 |
| Case 11, 55-60 y.o, M, 3T scanner | rectal | 0.68 | 0.18 | 11.9 | 8.8 | 5.9 | 0.304 |
| Case 12, 35-40 y.o, F, 3T scanner | breast | 0.34 | 0.23 | 13.3 | 8.6 | 5.9 | 0.254 |
| Case 13, 80-85 y.o, F, 3T scanner | endometrial | 0.46 | 0.31 | 9.9 | 9.8 | 5.8 | 0.263 |
| Case 14, 55-60 y.o, F, 3T scanner | skin cancer | 0.75 | 0.26 | 12.4 | 9.9 | 6.3 | 0.263 |
| Case 15, 55-60 y.o, M, 3T scanner | pancreatic | 0.83 | 0.18 | 12.0 | 8.5 | 5.4 | 0.270 |
| Case 16, 55-60 y.o, F, 3T scanner | breast | 0.78 | 0.15 | 10.6 | 8.5 | 4.8 | 0.397 |
| Case 17, 60-65 y.o, F, 1.5T scanner | endometrial | 0.41 | 0.27 | 9.9 | 8.7 | 5.5 | 0.346 |
| Case 18, 60-65 y.o, M, 1.5T scanner | esophagus | 0.003 | 0.04 | 6.7 | 7.0 | 4.5 | 0.234 |
| Case 19, 60-65 y.o, F, 1.5T scanner | skin cancer | 0.77 | 0.31 | 12.4 | 8.9 | 6.1 | 0.276 |
| Case 20, 50-55 y.o, F, 1.5T scanner | ovarian | 0.79 | 0.23 | 11.9 | 7.9 | 5.7 | 0.333 |
| Case 21, 65-70 y.o, M, 1.5T scanner | skin cancer | 0.71 | 0.37 | 13.2 | 8.6 | 7.2 | 0.144 |
| Case 22, 75-80 y.o, M, 1.5T scanner | skin cancer | 0.70 | 0.29 | 13.0 | 8.2 | 6.4 | 0.224 |
| Case 23, 65-70 y.o, M, 1.5T scanner | HCC | 0.40 | 0.24 | 12.3 | 8.6 | 6.2 | 0.274 |
| Case 24, 65-70 y.o, F, 1.5T scanner | gastric | 0.31 | 0.22 | 10.7 | 7.9 | 5.9 | 0.250 |
| Case 25, 60-65 y.o, F, 1.5T scanner | skin cancer | 0.55 | 0.23 | 13.2 | 8.4 | 6.0 | 0.299 |
| Case 26, 30-35 y.o, F, 1.5T scanner | breast | 0.73 | 0.25 | 12.0 | 8.2 | 5.5 | 0.382 |

**Table 5:** dMRI and histology metrics in cancer patients, with salient patients’ demographic and clinical information. The table reports dMRI metrics from fitting of an analytical, multi-compartment signal model of hindered EC diffusion and restricted IC diffusion within spherical cells. Patients’ age has been discretised with a granularity of 5-year intervals. HCC stands for hepatocellular carcinoma (primary liver cancer).

| Patient information / MRI scanner | Primary cancer | $f_{in\vert histo}$ | $f_{in\vert AN}$ | $vCS_{sph\vert histo}$<br>[$\mu m$] | $vCS_{cyl\vert histo}$<br>[$\mu m$] | $mCS_{histo}$<br>[$\mu m$] | $vCS_{AN}$<br>[$\mu m$] |
| --- | --- | --- | --- | --- | --- | --- | --- |
| Case 0, 75-80 y.o., F, 3T scanner | colon | 0.58 | 0.25 | 14.5 | 13.8 | 11.4 | 11.7 |
| Case 1, 60-65 y.o., M, 3T scanner | colon | 0.33 | 0.18 | 16.1 | 15.4 | 11.9 | 11.6 |
| Case 2, 60-65 y.o., M, 3T scanner | renal | 0.29 | 0.34 | 13.7 | 13.1 | 9.7 | 11.7 |
| Case 3, 65-70 y.o., M, 3T scanner | colon | 0.23 | 0.21 | 14.6 | 14.0 | 10.9 | 11.4 |
| Case 4, 65-70 y.o., M, 3T scanner | colon | 0.56 | 0.20 | 14.2 | 13.6 | 11.2 | 11.4 |
| Case 5, 70-75 y.o., M, 3T scanner | colon | 0.57 | 0.21 | 14.6 | 14.0 | 11.6 | 11.7 |
| Case 6, 65-70 y.o., M, 3T scanner | colon | 0.47 | 0.20 | 15.1 | 14.4 | 11.5 | 11.2 |
| Case 7, 45-50 y.o., F, 3T scanner | melanoma | 0.64 | 0.20 | 16.3 | 15.6 | 13.0 | 11.4 |
| Case 8, 75-80 y.o., F, 3T scanner | rectal | 0.57 | 0.12 | 14.1 | 13.6 | 11.4 | 11.5 |
| Case 9, 50-55 y.o., M, 3T scanner | HCC | 0.002 | 0.18 | 7.4 | 7.2 | 6.6 | 11.5 |
| Case 10, 60-65 y.o., F, 3T scanner | ovarian | 0.52 | 0.19 | 16.8 | 16.1 | 13.7 | 11.6 |
| Case 11, 55-60 y.o., M, 3T scanner | rectal | 0.68 | 0.17 | 14.7 | 14.1 | 11.9 | 11.7 |
| Case 12, 35-40 y.o., F, 3T scanner | breast | 0.34 | 0.25 | 16.1 | 15.5 | 13.3 | 11.4 |
| Case 13, 80-85 y.o., F, 3T scanner | endometrial | 0.46 | 0.29 | 14.5 | 13.6 | 9.9 | 11.4 |
| Case 14, 55-60 y.o., F, 3T scanner | melanoma | 0.75 | 0.24 | 15.1 | 14.5 | 12.4 | 11.4 |
| Case 15, 55-60 y.o., M, 3T scanner | pancreatic | 0.83 | 0.20 | 14.5 | 13.9 | 12.0 | 11.4 |
| Case 16, 55-60 y.o., F, 3T scanner | breast | 0.78 | 0.12 | 12.6 | 12.2 | 10.6 | 11.5 |
| Case 17, 60-65 y.o., F, 1.5T scanner | endometrial | 0.41 | 0.31 | 13.0 | 12.3 | 9.9 | 11.7 |
| Case 18, 60-65 y.o., M, 1.5T scanner | esophagus | 0.003 | 0.05 | 7.6 | 7.4 | 6.7 | 11.8 |
| Case 19, 60-65 y.o., F, 1.5T scanner | melanoma | 0.77 | 0.34 | 14.3 | 13.9 | 12.4 | 12.4 |
| Case 20, 50-55 y.o., F, 1.5T scanner | ovarian | 0.79 | 0.23 | 14.2 | 13.6 | 11.9 | 11.8 |
| Case 21, 65-70 y.o., M, 1.5T scanner | melanoma | 0.71 | 0.37 | 15.3 | 14.9 | 13.2 | 12.3 |
| Case 22, 75-80 y.o., M, 1.5T scanner | melanoma | 0.70 | 0.25 | 15.4 | 14.9 | 13.0 | 11.4 |
| Case 23, 65-70 y.o., M, 1.5T scanner | HCC | 0.40 | 0.25 | 15.5 | 14.9 | 12.3 | 12.2 |
| Case 24, 65-70 y.o., F, 1.5T scanner | gastric | 0.31 | 0.21 | 14.3 | 13.6 | 10.7 | 12.0 |
| Case 25, 60-65 y.o., F, 1.5T scanner | melanoma | 0.55 | 0.24 | 15.8 | 15.3 | 13.2 | 11.8 |
| Case 26, 30-35 y.o., F, 1.5T scanner | breast | 0.73 | 0.25 | 14.2 | 13.7 | 12.0 | 10.9 |

**Figure 10.**
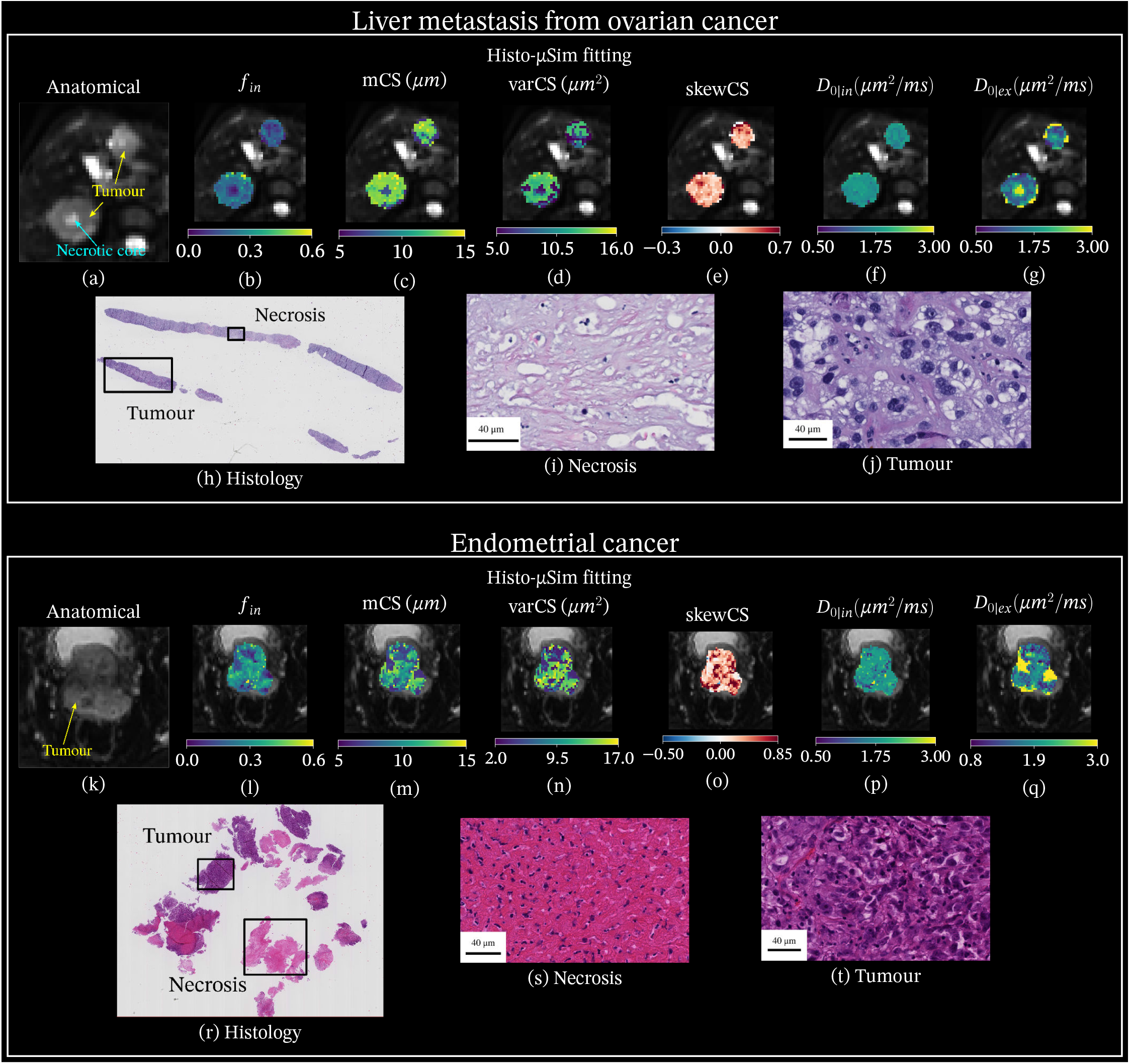
Parametric maps from *Histo-μSim forward model 2* as obtained on two representative patients *in vivo*, scanned on two different MRI scanners. Top: maps on ovarian cancer liver metastases (3T system); bottom: endometrial cancer (1.5T MRI system). From left to right, each panel reports a *b* = 0 image with the tumour outline (a and k) and then metrics *f*_*in*_ (b and l), mCS (c and m), varCS (d and n), skewCS (e and o), *D*_0|*in*_ (f and p) and *D*_0|*ex*_ (g and q). Below the metrics, details from a biopsy taken from one of the imaged tumours are also included (HE-stained biopsy in h and r; necrosis in i and s; active tumour in j and t).

## 3 Discussion

### 3.1 Summary and key findings

This article presents *Histo-μSim*, a new dMRI approach for microstructural parameter estimation informed by MC diffusion simulations within cellular environments reconstructed from histology. Our article has three main contributions. Firstly, it describes a practical step-by-step procedure, based entirely on freely available software, to reconstruct meshed cellular environments from histological images. These can be used to generate large dictionaries of realistic dMRI signals, coupled with histological properties. Secondly, it provides the scientific community with unique reference values of histology-derived cell size and density in non-cancerous and cancerous human liver tissues — information not easily found in the literature, yet essential to design the next-generation of cancer imaging techniques in radiology. Lastly, our paper showcases a numerical approach for dMRI parameter estimation informed directly by the simulated MC diffusion signals. The approach, feasible in cancer patients *in vivo*, is shown to outperform classical fitting of analytical signal models. As compared to the latter, *Histo-μSim* enables enhances parameter estimation on *in silico* data, and it deliver metrics that correlate more strongly with co-localised histology.

### 3.2 Simulation framework

Our simulation framework combines freely available software tools (i.e., QuPath [Bankhead et al., 2017], Inkscape and Blender) to reconstruct meshed cellular environments from 2D histological images. These are stored as sets of ASCII PLY files, a common file format for meshed geometrical models, being accepted by common open-source MC diffusion simulators such as MCDC [Rafael-Patino et al., 2020] or Camino [Hall and Alexander, 2009]. The procedure to convert histological data into PLY files has been described in detail in this article, and practical examples as well as tutorials for would-be users are provided in our freely accessible online repository, at the permanent address https://github.com/radiomicsgroup/dMRIMC. Our detailed guidelines equip the community with a practical tool to increase the realism of dMRI simulations, narrowing the gap between radiology and histology in cancer applications.

### 3.3 Substrates

To demonstrate our framework, we segmented 18 cellular environments from HE-stained liver tumour biopsies, referred to as *substrates*. These included tissues of different kinds, e.g., non-cancerous liver parenchyma as well as primary cancers of the liver and liver metastases, which were characterised in terms of cell density, IC area fraction, presence and morphology of EC luminal spaces, and CS distribution characteristics. We compiled a table reporting this information in a systematic manner, providing the community with reference histological values for cancer applications. To our knowledge, histology-derived cell morphometry literature has traditionally focussed on the study of neuroanatomy [Aboitiz et al., 1992, Duval et al., 2019], and limited quantitative data are available in body tissues or cancer, especially in relation to CS. Information on the expected CS and cell denisity of a tissue is essential to optimise dMRI acquisition protocols, e.g., to design b-values or diffusion times. Therefore, delivering such a data base is a major contribution of our work, as it may be used to devise innovative dMRI acquisition protocols tailored for body imaging.

#### 3.4 Simulation-informed parameter inference *in silico*

We investigated whether synthetic dMRI signals generated through our histology-informed framework can be used to devise new strategies for CS distribution mapping, urgently sought for cell population profiling in oncology [Jiang et al., 2020a, Hoffmann et al., 2023]. To this end, we interpolated the discrete dictionary of paired examples of tissue parameters and synthetic dMRI signals using Radial Basis Function (RBF) regressors. This provided numerical forward models that do not rely on approximated analytical functional forms for the IC/EC signal, e.g., restricted diffusion within cells of regular shape and equal size [Panagiotaki et al., 2015, Jiang et al., 2020b], or Gaussian EC diffusion. Such forward models can be easily embedded into routine non-linear least squares (NNLS) fitting, based on likelihood maximisation [Panagiotaki et al., 2012].

We compared the performance of our approach in predicting a single CS (effective cell diameter) statistic [Veraart et al., 2020] against standard analytical approaches based on restricted diffusion within cylinders. Results not only point towards the superiority of our approach in CS estimation, but also show benefit in the estimation of other diffusion properties, such as the intrinsic cytosolic diffusivity or the IC fraction. We also studied the feasibility of estimating the intrinsic EC diffusion coefficient and the first three CS distribution moments, without imposing any analytical functional form to the CS distribution (forward model 2). We observe satisfactory performances in the estimation of *D*_0|*ex*_, mCS and varCS, and weaker performances for skewCS (moderate correlation between ground truth and estimated values for *D*_0|*ex*_; strong to very strong correlations for mCS; moderate to strong for varCS). Overall, these findings demonstrate that our proposed approach can be deployed for geometries and dMRI sequences for which analytical expressions may not be readily available, provided that they can be simulated, paving the way to numerical, equation-free dMRI models with enhanced biological specificity.

### 3.5 Simulation-informed parameter inference in fixed *ex vivo* mouse tissue

After demonstrating CS distribution mapping *in silico*, we tested whether it is also feasible on actual MRI scans. For this experiment, we analysed both pre-clinical *ex vivo* data from 8 mouse tissue samples, as well as *in vivo* scans acquired on cancer patients with two clinical MRI systems. Notably, the tissue scanned on the pre-clinical system was considerably different from that used to build the numerical signal models (e.g., mouse breast tumours, kidneys, and spleens, versus human liver parenchyma and liver tumours), and thus served as useful *out-of-distribution* test bed for generalisation. The parametric maps obtained *ex vivo* show a number of interesting and potentially relevant inter-sample and intra-sample contrasts, which are in most cases confirmed by histology both qualitatively and quantitatively. The co-localised MRI and histology data acquired in mice enabled a detailed MRI-correlation analysis, which essentially confirms findings from *in silico* experiments. We observed a strong correlation between dMRI and histological *f*_*in*_ and mCS; a moderate, positive correlation for varCS; and a weak, positive correlation for skewCS. These correlation demonstrate the potential of *Histo-μSim* to boost the biological specificity of dMRI towards cancer, and are encouraging, given i) the relatively small size of our sample; ii) the inherent difficulty of ensuring accurate co-localisation between dMRI and histology; iii) the differences between the substrates used to build the models and the tissue imaged *ex vivo*; iv) the fact that these MRI-histology correlations were stronger than those from standard analytical signal model. All in all, the *ex vivo* experiments suggest that *Histo-μSim* may provide new biomarkers of tissue microstructure that may shed new light onto the presence of different cell populations in a voxel, through CS distribution mapping.

### 3.6 Simulation-informed parameter inference in cancer patients *in vivo*

Lastly, we demonstrated *Histo-μSim* in a pilot cohort of patients *in vivo*, and compared *Histo-μSim* metrics to histological indices from HE biopsies collected from one of the imaged tumours. This final demonstration shows that *Histo-μSim* maps can be obtained with dMRI scans that are feasible in the clinic, i.e., not exceeding 15 minutes, with moderate maximum b-values (around 1500 s/mm_2_), and based on vendor-provided sequences. The inspection of parameteric maps reveals key inter-tumour and intra-tumour contrasts, which are plausible given the high microstructural heterogenity seen in the HE-stained biopsied tissue. For example, areas lying within tumour necrotic cores show reduced *f*_*in*_ and mCS, compatible with necrosis and presence cell debris.

The collection of biopsy data enabled a second MRI-histology correlation study. Despite the inherent challenge of relating a small sliver of biopsied tissue to MRI metrics evaluated over large tumours, the new biopsy-MRI comparison confirms that *Histo-μSim* provide metrics that correlate more strongly to their histological counterparts than standard analytical signal models. This results suggests, again, that *Histo-μSim* may contribute to increasing the biological specificity of dMRI, compared to current state-of-the-art multi-exponential approaches. Nevertheless, we acknowledge that in this case correlations between dMRI and histology are weaker. The observed correlation levels are not surprising given that we could not locate the exact tumour location where the needle was inserted. Because of this, we included all MRI voxels within the tumour to obtain per-tumour MRI metrics in our MRI-histology comparison, a fact that has reduced the accuracy of the co-localisation between the two modalities. Nevertheless, we also acknowledge that other factors may have contributed to explaining the difference in correlation seen on *in vivo* human data, compared to *ex vivo* mouse tissue. These could include, for example, the fact that this first demonstration of *Histo-μSim* does not account for IC/EC water exchange, a phenomenon that is likely to play a larger role *in vivo*, due to active transcytolemmal water transport [Li et al., 2017].

### 3.7 Methodological considerations and limitations

We acknowledge some potential limitations of our approach. The first one relates to the manual reconstruction of virtual tissue environments form histology. Despite some remaining inaccuracies, the manual outlining has enabled the accurate segmentation of cell boundaries, difficult to achieve with high accuracy and high precision through automatic cell segmentation software such as QuPath [Bankhead et al., 2017] (Supplementary Table S1). Nevertheless, we acknowledge that the approach is inherently slow and difficult to scale up to create large dictionaries of synthetic signals and histological properties, essential to support advanced parameter estimation techniques (e.g., through deep learning). In future, we plan to expand our tissue environment data bases through automatic histological image processing, and explore more sophisticated parameter estimation methods as those used in the first demonstration of *Histo-μSim*.

Secondly, we built virtual tissue environments effectively characterised by cylindrical geometries, and then focussed on the analysis of 2D diffusion. This was due to the availability of a large data set of HE-stained sections in human and mouse tissue (inherently 2D). In future, we plan to perform simulations that capture the full 3D complexity of the tissue substrates, reconstructing these, for example, from 3D micrographs [Lee et al., 2020b] or from 3D confocal microscopy [Khan et al., 2015] data.

Thirdly, in this study we illustrated the benefits of relaxing some of the constraints and hypotheses underlying standard analytical diffusion models through numerical simulations. However, we point out that the first demonstration is not free from assumptions, since *Histo-μSim* tissue parameter estimates inherit the hypotheses made to conduct the MC simulations themselves. It is possible that some of discrepancies observed between dMRI and histology may have been exacerbated by important microstructural properties that were not accounted for in our simulated random walks, e.g., transcytolemmal water exchange [Jiang et al., 2022]. Unaccounted exchange can bias the estimation of the IC fraction *f*_*in*_ [Gardier et al., 2023], a fact that may contribute to explaining, for example, the dMRI underestimation of *f*_*in*_ compared to histology in the fixed mouse kidneys, or in patients *in vivo*. In future, we plan to increase the complexity of our MC simulations, and account for important aspects of tissue microstructure so far neglected, including water exchange and beyond, e.g.: variability in intrinsic diffusivity among cells or between lumina and EC space; differences in intra-compartament relaxation properties [Lemberskiy et al., 2018, Fieremans and Lee, 2018, Palombo et al., 2023]. Related to this point, we also acknowledge that our signal models do not account for contributions coming from intra-voxel incoherent flow within capillaries [Le Bihan et al., 1986], which we did not simulate. For this reason, we took care to exclude b > 100 *s*/*mm*^2^ measurements *in vivo*, where vascular signals are negligible [Cui et al., 2015]. In future, we aim to increase the realism of our simulations by including a third compartment of capillary perfusion, alongside IC and EC diffusion.

Additionally, when analysing histological images for MRI-histology validation, we segmented cellular structures manually for the *ex vivo* mouse data, while we used the automated cell segmentation for the analysis of patients’ biopsies. We did not carry out manual cell segmentation for the patients’ data because it was not possible to identify the exact within tumour location on dMR images from which the biopsy was taken. Due to this, metrics from all tumour tissue found on the HE had to be compared to a whole tumour seen on dMRI, making manual cell segmentation on HE images unfeasible. Comparisons between manual and automatic QuPath [Bankhead et al., 2017] cell segmentation show that while QuPath-derived varCS and skewCS differ considerably from varCS and skewCS from manual segmentations (high bias for the former, poor correlation for the latter), QuPath-derived vCS and mCS are acceptable surrogates of their manually-derived counterparts (Supplementary Table S1). For this reason, we only included mCS and vCS in the biopsy-MRI correlation analysis *in vivo*, excluding varCS and skewCS. Future work will address this limitation through optimised pipelines for automatic CS distribution moments computation on HE histology.

Lastly, in this work we did not study advanced diffusion encodings such as oscillating gradients [Jiang et al., 2020b], double diffusion [Shemesh et al., 2016] or *b*-tensor [Westin et al., 2016] encoding, since we focussed on off-the-shelf, widespread clinical protocols. In future we aim to simulate more advanced dMRI acquisition, beyond routine PGSE. These may give access to more detailed information on cancer microstructure than standard diffusion encoding, and potentially improve the estimation of promising metrics such as skewCS, with important applications in non-invasive cell profiling in cancer [Hoffmann et al., 2023].

## 4 Conclusions

*Histo-μSim*, a new dMRI parameter estimation approach informed by MC simulations within tissue environments reconstructed from histology, provides histologically-meaningful indices within clinically-acceptable scan times. The method outperforms standard multi-compartment analytic models on *in silico* data, as well as in dMRI scans acquired on fixed mouse tissue *ex vivo* and on cancer patients *in vivo. Histo-μSim* may therefore play a key role in the development of new essays for the non-invasive characterisation of solid tumors in the body, and thus contribute to bringing precision oncology one step closer to the clinic.

## 5 Materials and Methods

### 5.1 Simulation framework

In our framework, illustrated in Fig. 1, we create 3D meshes of histological structures, such as cells, from segmentations drawn on histological images. These meshes can be used to generate random walks in MC simulations and, finally, dMRI signals, for any dMRI protocol of interest. We proceed as follows.

First, a histological image is opened with QuPath [Bankhead et al., 2017] and a ROI is selected and cropped, taking care to include in the image the scale of magnification. The image is then opened in Inkscape, where cells and other geometric features are manually segmented and separated into layers. We segmented cells and cell debris, luminal spaces, and vessels. Here we demonstrate the framework with careful, manual segmentation, but automatic segmentations would also be possible. Two types of files are then exported: a 3D object with all the features included included in a single SVG file, as well as an individual SVG file for each feature. The SVG format is used as it allows for further manipulation with Blender. In Blender, SVG files are then transformed into 3D ASCII PLY triangular meshes. We reconstructed 2D cellular environments from standard HE biopsies and obtained 3D meshes by simply replicating 2D contours along the trough-plane direction, thus generating cylinders with irregular sections. Nonetheless, 3D segmentations could also be used (e.g., from 3D confocal microscopy).

Meshes are fed to the MCDC Simulator, an open-source MC engine [Rafael-Patino et al., 2020], in order to synthesise water molecules Brownian random walks within the substrate. We used the large mesh including all the segmented structures to generate EC random walks, by seeding walkers outside all such structures, uniformly. Conversely, we generated IC random walks for each cell independently. For this first proof-of-concept we did not model permeability of barriers, i.e., we did not account for exchange between EC and IC spaces, with walkers experiencing elastic collisions at cell boundaries.

We used 10 linearly-spaced values in the range [0.8, 3] *μm*^2^/*ms* for both IC and EC intrinsic diffusivities (referred to as *D*_0|*in*_ and *D*_0|*ex*_), covering all possible combinations of the two (100 unique (*D*_0|*in*_, *D*_0|*ex*_) pairs for each substrate). Each simulation was run with a duration of *T* = 140 *ms* and step number of *N*_*step*_ = 3000. The IC simulations were performed with 1000 water molecules for each cell, while EC random walks were obtained for 10 000 walkers. As mentioned, vessel structures were included in the segmentation as they influence the patterns of EC diffusion, but they were not seeded with walkers (so they do not directly generate MRI signal). Regarding lumen spaces, spin walks were generated similarly to the IC case: spins were seeded within each lumen independently, and no crossing of the lumen membrane was possible (elastic reflection in presence of collisions; 1000 walkers per lumen).

Lastly, custom-written python code was used to synthesise dMRI signals from the random walks for a given acquisition protocol of interest. The signal *s*_*n*_ for the *n*-th structure is [Fieremans and Lee, 2018]

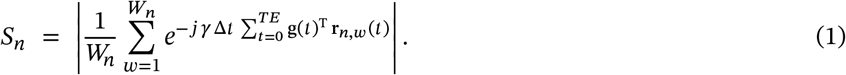

Above, **r**_*n*,*w*_(*t*) is the *w*-th walker trajectory within the *n*-th structure; Δ*t* = *T*/*N*_*step*_ is the temporal resolution; *T* is the simulation duration; and **g**(*t*) is the diffusion-encoding gradient. The IC/EC signals *S*_*in*_/*S*_*ex*_ were respectively obtained as

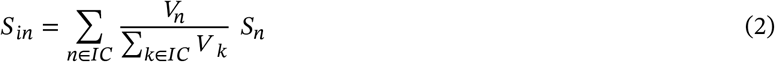

and

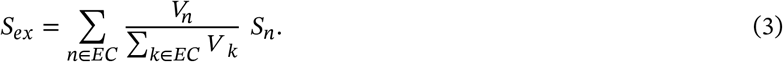

with *V*_*n*_/*V*_*k*_ standing for the volume of the *n*-th/*k*-th structure. The index *n* loops over cells (including debris) in Eq. 2, referring to the IC signal; likewise, it loops over the set of EC structures in Eq. 3, i.e., the EC space itself and the luminal spaces. Last, the total signal is

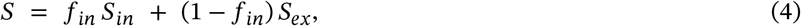

with *f*_*in*_ being the IC volume fraction. Note that signal fractions *f*_*in*_ and *f*_*ex*_ = 1 − *f*_*in*_ are *T*_2_-weighted in principle, given that the IC/EC spaces may feature different *T*_2_ constants [Lemberskiy et al., 2018, Palombo et al., 2023]. Nonetheless, in this first demonstration of our MC framework, we do not account for intra-compartment relaxation properties, in order to reduce the number of tissue parameters required to characterise the signal.

A repository with step-by-step guidelines on how to implement the framework is released at https://github.com/radiomicsgroup/dMRIMC.

### 5.2 Reconstruction of virtual tissue environments

We reconstructed 18 cellular environments, referred to as *substrates* from now on. These were derived from biopsies of malignant solid tumours of the liver (primary cancer and metastatic) of 10 different patients (1 to 3 substrates drawn per patient, see Table 1), acquired as part of ongoing imaging studies at the Vall d’Hebron Institute of Oncology (Barcelona, Spain). The substrates spanned a rich set of different cytoarchitectures, from non-cancerous liver parenchyma to cancer areas, such as dense cancer cell packings, fibrosis, necrosis, and a mix of all the above.

We characterised each substrate with the following microstructural parameters:

- ROI area and cellularity (number of cells per *mm*^2^ of biopsied tissue);
- IC area fraction *f*_*in*_;
- lumen fraction of EC area *f*_*l*_;
- lumen diameters 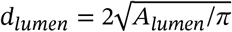, with *A*_*lumen*_ being the segmented lumen area;
- mean CS index mCS = ⟨*d*_*cell*_ ⟩, where 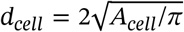 is the individual cell diameter calculated from its area *A*_*cell*_, and ⟨… ⟩ is the average over the distribution in a substrate;
- CS variance index varCS = ⟨(*d*_*cell*_ − mCS)^2^⟩;
- CS skewness index

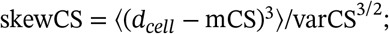
- volume-weighted CS (vCS) index for a system with spherical geometry [Novikov et al., 2019, Grussu et al., 2022] (vCS_*sph*_), defined as

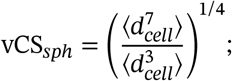
- vCS index for a system with cylindrical geometry [Burcaw et al., 2015, Veraart et al., 2020] (vCS_*cyl*_), defined as

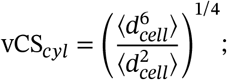
- shape *h* (dimensionless) and scale *c* (units: *μm*) parameters of a gamma-distribution [Assaf et al., 2008]

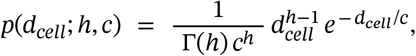

fitted to the set of cell diameters {*d*_c*ell*,1_, *d*_c*ell*,2_, …}. Above, 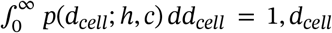 is the generic cell diameter (units: *μm*), and Γ(*z*) is the Gamma function

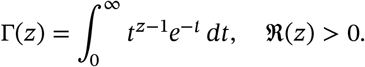

### 5.3 In silico assessment of the dependence of IC diffusion on CS distribution moments

We investigated whether salient features of the underlying CS distribution are encoded in the dMRI signal for realistic, clinically-viable acquisitions. We studied the relationship between the IC ADC and mean, variance and skewness of the CS distribution (mCS, varCS, and skewCS), calculating Pearson’s correlation coefficients between ADC_*in*_ and each of this CS indices in turn. For ADC_*in*_ estimation, we synthesised dMRI signals using *D*_0|*in*_ = 2.2 *μm*^2^/*ms* for b = {0, 100, 200, 300, 400, 500} *s*/*mm*^2^, averaging magnitude signals from two orthogonal gradient directions for each b, and perpendicular to the substrate longitudinal axis. We then estimated ADC_*in*_ by regressing ln(*S*_*in*_) = −*b* ADC_*in*_, fixing *δ* = 15 ms and varying Δ (Δ ∈ {15, 25, 35, 45, 55, 65} *ms*).

### 5.4 Development of simulation-informed parameter inference

We investigated the potential utility of our synthetic signals to inform microstructural parameter estimation. We synthesised DW signals according to three dMRI protocols, matching those implemented for the acquisition of *ex vivo* and *in vivo* MRI data (see sections below). We simulated 100 signals per substrate (10 values of *D*_0|*in*_ × 10 values of *D*_0|*ex*_), for a total of 1800 signals. For all protocols, we obtained the final measurement set by averaging signals generated for two orthogonal directions, perpendicular to the substrate longitudinal axis. The protocols were:

- **PGSE-in**: a PGSE protocol, matching that implemented on a 3T clinical system *in vivo*. It consisted of 3 b = 0 and 18 DW measurements, namely: b = {50, 100, 400, 900, 1200, 1500, 50, 100, 400, 900, 1200, 1500, 50, 100, 400, 900, 1200, 1500} *s*/*mm*^2^, *δ* = {3.9, 5.2, 9.2, 15.0, 18.2, 21.0, 3.9, 5.2, 9.2, 13.0, 15.8, 18.5, 3.9, 5.2, 9.2, 13.0, 15.8, 18.5} *ms*, Δ = {27.8, 29.0, 33.0, 28.7, 31.8, 34.7, 7.8, 29.0, 33.0, 37.0, 39.6, 42.3, 7.8, 29.0, 33.0, 37.0, 39.6, 42.3} *ms*.
- **TRSE**: a DW twice-refocussed spin echo (TRSE) protocol, matching that implemented on a 1.5T clinical system *in vivo*. It consisted of 3 b = 0 and 18 DW measurements, namely: b = {0, 50, 100, 400, 900, 1200, 1600} *s*/*mm*^2^, repeated for 3 different diffusion times. The duration/separation of the gradient lobes (Supplementary Figure 1) for the 3 diffusion times were: *δ*_1_= {8.9, 13.2, 18.9} *ms, δ*_2_= {17.6, 19.3, 21.0} *ms, δ*_3_ = {20.4, 24.8, 30.5} *ms, δ*_4_ = {6.0, 7.7, 9.5} *ms*, Δ_1,2_ = {17.4, 21.7, 27.5} *ms*, Δ_1,4_ = {63.9, 74.2, 87.5} *ms*.
- **PGSE-ex**: a second PGSE protocol, matching that implemented on a pre-clinical 9.4T system for *ex vivo* imaging. It consisted of 2 b = 0 and 6 DW measurements, namely: b = {0, 500, 2000, 4500} *s*/*mm*^2^ acquired for each of Δ = {16.5, 37.0} *ms*, with *δ* = 12 *ms*.

We then interpolated the set of paired examples of tissue parameters **p** and dMRI signals *s*(**p**) with a RBF regressor, which implements the forward model **p** ↦ *s*(**p**). This was finally embedded into routine NNLS fitting, based on maximum-likelihood estimation [Panagiotaki et al., 2012]. To test the feasibility of using simulation-informed forward models for parameter estimation, we performed a leave-one-out experiment. Briefly, for all substrates in turn, we learnt **p** ↦ *s*(**p**) on noise-free signals from 17/18 substrates, and then plugged the learnt model in NNLS fitting of the noisy signals from the 18^th^ substrate (Rician noise; *b* = 0 signal-to-noise ratio (SNR) of 50).

We performed fitting twice, considering two different forward models **p** ↦ *s*(**p**):

- in *forward model 1*,

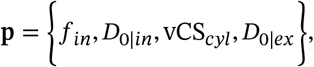

estimating a single CS statistic (vCS_*cyl*_) per voxel. We chose to estimate vCS_*cyl*_, rather than mCS, to enable the comparison of model 1 to fitting a well-established multi-compartment analytical signal model (see below);
- in *forward model 2* instead,

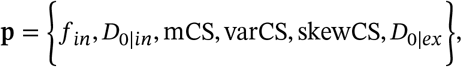

estimating the first three moments of the full CS distribution.

The quality of parameter estimation was assessed by scatter density plots and Pearson’s correlation coefficients between estimated and ground truth parameter values. Moreover, fitting of forward model 1 was compared to a widely-used analytical model, describing the dMRI signal as the sum of IC/EC contributions, i.e.,

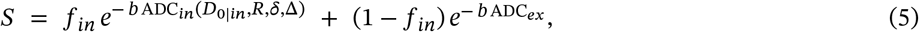

where ADC_*in*_ characterises signal decay due to restriction within cells. This approach is used, for example, in popular techniques such as VERDICT and IMPULSED [Panagiotaki et al., 2015, Jiang et al., 2020b]. However, while VERDICT and IMPULSED ADC_*in*_ is based on a model of spherical cells, here we used the expression for diffusion within cylinders, given the cylindrical symmetry of our substrates. We used an effective radius 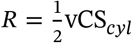. We used vCS_cyl_ rather than mCS since the former accounts for the fact that larger cells contain more water, and hence contribute more to the DW signal, than smaller cells [Veraart et al., 2020]. Nontheless, we point out that vCS_*cyl*_ is a metric prone to mesoscopic fluctuations, being highly sensitive to the tails of the cell size distribution within a voxel, with increasing sensitivity the smaller the voxel gets [Novikov et al., 2019]. In practice, ADC_*in*_ in Eq. 5 is written as

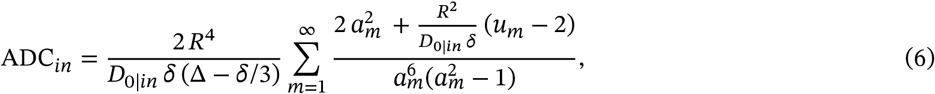

where

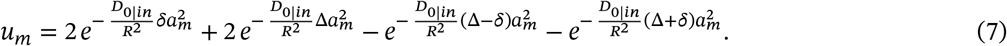

Above, *D*_0|*in*_ is the intra-cylinder diffusivity, *a*_*m*_ is the m-th root of *J*_1_(*x*) = 0, with *J*_1_(*x*) being the Bessel function of the first kind, order 1, and 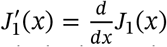 [P. Vangelderen et al., 1994]. Note that the analytical model in Eq. 6, while common in dMRI literature, lacks higher-order terms in each compartment. It is therefore only a crude approximation of the signal at a given *b, δ* and Δ — this fact is indeed what motivates our efforts to build numerical dMRI signal models informed directly by simulations, which do not rely on approximated analytical signal expressions.

Both MC simulation-informed and analytical signal model fitting were performed with the freely-available *bodymritools* python tools (https://github.com/fragrussu/bodymritools; scripts *mri2micro _dictml*.*py* and *pgse2cylperpinex*.*py*).

#### Histological validation of *Histo-μSim* MRI on *ex vivo* mouse tissue

We tested simulation-informed fitting on pre-clinical dMRI data, which were acquired on 8 formalin-fixed *ex vivo* mouse tissue specimens, namely: a non-cancerous breast sample; 3 breast tumours from the MMTV PyMT model [Guy et al., 1992, Attalla et al., 2021], obtained at weeks 9, 11 and 14; a normal spleen and a spleen suffering from splenomegaly, secondary to advanced breast cancer in one MMTV mouse; two kidneys from C57BL/6 WT male mice (9 weeks old), one normal and one with folic acid-induced injury [Yan, 2021]. Mice were housed at the Specific Pathogen-Free barrier area of the Vall d’Hebron Institute of Oncology (VHIO). All animal procedures were approved by the Animal Care unit and the Ethics Committee for Animal Experimentation (CEEA) of the Vall d’Hebron Research Institute (VHIR) and the Generalitat de Catalunya, and were performed according to the European legal framework for research animal use and bioethics. Animals were monitored daily and euthanised upon signs of humane endpoints. Two mouse models were used, generating breast, spleen and kidney samples. These were processed for further histological analyses, as part of ongoing studies at VHIO. A dMRI scan of the tissue was performed at room temperature before inclusion in paraffin for histology.

#### MMTV-PyMT transgenic mouse model

The MMTV-PyMT FVB/NJ mouse strain [Guy et al., 1992] is commonly employed to mimic human breast cancer progression [Attalla et al., 2021]. The model relies on the MMTV long terminal repeat promoter, which drives the expression of the antigen of PyMT, a potent oncogene. These transgenic mice are viable despite loss of lactational ability, which is coincident with the transgene expression. Breast tumours arise in virgin and breeder females as well as in males starting from 9 weeks of age. Splenomegaly is also observed at the latter stages of the tumour growth. For this study, we used 4 MMTV-PyMT FVB/NJ female mice, which were euthanised by CO_2_ asphyxiation at different time points to collect the following samples: non-cancerous breast and non-pathological spleen (2 weeks); a breast tumour at weeks 9, 11 and 14; an enlarged spleen (splenomegaly) at late stage cancer (14 weeks).

#### Folic acid-induced kidney injury

The folic acid-induced kidney injury mouse model is based on the fact that high doses of folic acid are toxic, despite being the same substance beneficial at low doses [Yan, 2021]. For this study, we used two male mice (C57BL/6 WT, approximately 9 weeks old), which were intra-peritoneally injected with a single dose of vehicle (300 mM NaHCO_3_) or with folic acid (250 mg/kg). 30 days after the injection, mice were euthanised by CO_2_ asphyxiation and the kidneys were collected for downstream processing.

#### dMRI acquisition

Briefly, collected tissues were fixed for 24 hours in buffered 4% formaldehyde, transferred to phosphate-buffered saline (PBS) solution and embedded in 1% agarose gel dissolved in PBS, within a histological cassette. Embedded samples were kept in PBS solution, and scanned at room temperature on a 9.4T Bruker Avance system, with 200 *mT*/*m* gradient insert and a RX/TX birdcage coil. The protocol included a high resolution anatomical T2-weighted RARE scan, and dMRI (DW spin echo), with the protocol matching the PGSE-ex protocol described above (see Materials and Methods; same nominal b-values, and same gradient timings). Other salient dMRI scan parameters were: fat suppression with a frequency-selective 90 degree gauss512 pulse (bandwidth: 1400.1 Hz); resolution 0.2 × 0.2 × 0.57 *mm*^3^, TE = 55.1 *ms*, TR = 2250 *ms*, 3 mutually-orthogonal direction for each gradient timing and b-value.

#### Histology acquisition

After MRI, samples were transferred to 70% ethanol for 24 hours and then embedded in paraffin. 3-*μm* sections were obtained on a manual microtome and stained with HE, using a robust carousel tissue stainer (Slee Medical) according to common methods. Digital images of the HE-stained sections were acquired on a Hamamatsu C9600-12 scanner (resolution: 0.45 *μm*).

#### dMR image processing

dMRI scans were denoised [Veraart et al., 2016] and Gibbs ringing was mitigated [Kellner et al., 2016]. Maps from forward model 2 were computed voxel-by-voxel, via NNLS regularised maximum-likelihood fitting. Metrics were: *f*_*in*_, *D*_0|*in*_, mCS, varCS, skew When learning the forward signal model via RBF regression, we pooled together all 1800 signals from all substrates. For comparison, we also fitted an analytical signal model voxel-by-voxel. The model accounted again for restricted IC diffusion and hindered EC diffusion, and is thus equivalent to that of Eq. 5. However, in this case we used the expression of IC ADC derived for diffusion within spheres, rather than for cylinders [Balinov et al., 1993]. For all model fitting (MC-informed and analytical), L2 regularisation of the fitting objective function was used. The freely-available *bodymritools* python tools were used (https://github.com/fragrussu/bodymritools; scripts *mri2micro _dictml*.*py* and *pgse2sphereinex*.*py*)

We computed mean and standard deviation of IC fraction and CS statistics in 18 ROIs, drawn in homogenous areas, far from edges and from the location of sharp contrasts on the *b* = 0 dMRI image. We indicated the metrics as follows: *f*_*in*|*MC*_, mCS_*MC*_, varCS_*MC*_ and skewCS_*MC*_ for *Histo-μSim* MC-informed fitting; *f*_*in*|*AN*_ and vCS_*sph*|*AN*_ for analytical model fitting.

#### Histological image processing

In parallel, we also processed the HE images to obtain histological counterparts of MRI metrics. We manually segmented cells in areas whose location visually matched the position of the dMRI ROIs, and computed *f*_*in*|*histo*_, vCS_*sph*|*histo*_, vCS_*cyl*|*histo*_, mCS_*histo*_, varCS_*histo*_, skewCS_*histo*_ given the set of segmented cells, as illustrated for the tissue environment generation above. Segmentation was not performed in areas rich of fat as seen on HE images, given that dMRI acquisitions are fat-suppressed. For reference, we also obtained cell segmentations automatically, using QuPath [Bankhead et al., 2017], and compared manually-derived and QuPath-derived histological metrics. For this, we calculated Pearson’s correlations between manually-derived and QuPath-derived histological metrics. Additionally, we also evaluated a Bias Index (BI), defined as *BI* = *median*(*E*), where 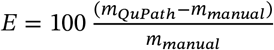 represent the generic metric obtained from QuPath/manual segmentations, and IQR is the inter-quartile range. BI quantifies the bias of QuPath-derived histological indices compared to reference values obtained through manual cell segmentation.

#### MRI-histology correlation analysis

We computed Pearson’s correlation coefficients among all possible pairs of MRI and histological metrics, namely: *f*_*in*|*histo*_, vCS_*sph*|*histo*_, vCS_*cyl*|*histo*_, mCS_*histo*_, varCS_*histo*_ and skewCS_*histo*_ (manually-derived histological metrics); *f*_*in*|*MC*_, mCS_*MC*_, varCS_*MC*_, skewCS_*MC*_, *D*_0|*in*|*MC*_ and *D*_0|*ex*|*MC*_ (*Histo-μSim* MC-informed fitting); *f*_*in*|*AN*_, vCS_*sph*|*AN*_, *D*_0|*in*|*AN*_ and *ADC*_*ex*|*AN*_ (analytical model fitting).

### 5.5 In vivo demonstration of Histo-μSim in cancer patients

Lastly, we also tested the feasibility of *Histo-μSim* in actual *in vivo* dMRI scans of cancer patients, which were acquired as part of ongoing imaging studies at the Vall d’Hebron Institute of Oncology. All participants provided informed written consent, and were scanned in imaging sessions approved by the Clinical Research Ethics Committee (CEIm) of the Barcelona Vall d’Hebron University Hospital (VHUH) (code: PR(AG)29/2020)). We studied scans from 27 patients suffering from advanced solid tumours, candidate for phase I clinical trials at the VHUH, and referred to as Cases 0 to 26, with the case ID being randomly generated for this article. Scans were acquired at either abdominal or pelvic level, using a 1.5T Siemens Avanto system (10 cases) and a 3T GE SIGNA Pioneer system were used (17 cases). Additionally, a biopsy was collected from one of the tumours approximately one week after MRI.

#### dMRI acquisition

For the 1.5T system, the protocol included anatomical T2-weighted fast spin echo imaging and dMRI. dMRI was based on a DW TRSE Echo Planar Imaging (EPI) sequence, with the diffusion encoding protocol matching exactly the TRSE protocol used in simulations (see above for details). Other salient parameters were: resolution 1.9 × 1.9 × 6 *mm*^3^, TE = {93, 105, 120} *ms*, TR = 7900 *ms*, trace DW imaging, NEX = 2, GRAPPA = 2, 6/8 Partial Fourier imaging, BW = 1430 Hz/pixel, acquisition of a *b* = 0 image with reversed phase encoding. For the 3T scanner instead, the protocol also included anatomical T2-weighted fast spin echo imaging and dMRI, acquired with PGSE EPI according to the “PGSE-in” protocol described in simulations above. Other salient parameters were: resolution 2.4 × 2.4 × 6 *mm*^3^, TE = {75, 90, 105} *ms*, TR ≈ 3000 *ms* (respiratory gated), trace DW imaging, NEX = 2, ASSET = 2, BW = 1953 Hz/pixel. The dMRI protocol took approximately 15 minutes in both machines. A schematic of the PGSE and TRSE DW sequences is included in Supplementary Fig. S1.

#### Histology acquisition

We obtained ultrasound-guided biopsies from one of the imaged tumours, obtained approximately one week after dMRI. The histological material underwent standard processing, form which we obtained HE-stained sections, which we digitised a Hamamatsu C9600-12 slide scanner (resolution: 0.45 *μm*).

#### dMR image processing

Scans were denoised [Veraart et al., 2016], corrected for Gibbs ringing [Kellner et al., 2016] and motion, and EPI distortions mitigated (1.5T system only) [Andersson et al., 2003]. Afterwards, each DW image was normalised to the *b* = 0 acquired at the corresponding TE [Panagiotaki et al., 2015], and *forward model 2* was fitted voxel-by-voxel (regularised maximum-likelihood NNLS fitting; images for *b* ≤ 100 *s*/*mm*^2^ were excluded to minimising vascular contributions) within tumours, whose outline was drawn manually on the dMRI scan by an experienced radiologist (R.P.L.). For comparison, we also fitted the same multi-exponential analytical model deployed on the *ex vivo* mouse scans, accounting for restricted IC diffusion within spherical cells and hindered extra-cellular diffusion. Note that to our knowledge, no analytical signal expression exists for restricted IC diffusion within spherical cells for the TRSE acquisition. For this reason, we replaced the IC analytical signal expression with a continuous RBF interpolation of signals generated through MC simulations [Rafael-Patino et al., 2020] within meshed spheres. For all model fitting (MC-informed and analytical), L2 regularisation of the fitting objective function was used.

Finally, mean values of all dMRI metrics within the tumours were extracted.

#### Histology image processing

An experienced pathologist (S.S.) inspected HE-stained biopsies and outlined the tumour tissue, on which we segmented cells automatically using QuPath [Bankhead et al., 2017]. Segmented cells were collected and per-biopsy histological metrics were computed.

#### MRI-histology correlation analysis

Similarly to what was performed with mouse dMRI data, we evaluated Pearson’s correlation coefficients between histological and dMRI metrics. To this end, we obtained per-patient representative dMRI indices by averaging parametric maps across tumoural voxels. In summary, we focussed on the following metrics. For histology: IC fraction *f*_*in*|*histo*_, volume-weighted CS and mean CS (vCS_*cyl*|*histo*_, vCS_*sph*|*histo*_ and mCS_*histo*_). For *Histo-μSim*: IC fraction *f*_*in*|*MC*_ and mCS_*MC*_. For the analytical signal model: IC fraction *f*_*in*|*MC*_ and vCS_*sph*|*AN*_. Note that in histology we did not compute varCS and skewCS since QuPath does not allow for their reliable estimation (Supplementary Table S1).

## Data Availability

The Monte Carlo simulation framework upon which Histo-microSim has been developed is freely available in GitHub at the permanent address: https://github.com/radiomicsgroup/dMRIMC. The virtual tissue environments reconstructed from histology and the mouse MRI and histology data have been released publicly. The final, permanent addresses of the data bases can be found on the GitHub repository (link: https://github.com/radiomicsgroup/dMRIMC). The data in mouse tissue and in cancer patients used for all MRI-histology correlation analyses are included as tables in this manuscript. Routines for simulation-informed fitting are freely available as part of BodyMRITools at the permanent address: https://github.com/fragrussu/bodymritools (script mri2micro_dictml.py).

## Acknowledgments

VHIO would like to acknowledge: the State Agency for Research (Agencia Estatal de Investigación) for the financial support as a Center of Excellence Severo Ochoa (CEX2020-001024-S / AEI / 10.13039 / 501100011033), the Cellex Foundation for providing research facilities and equipment and the CERCA Programme from the Generalitat de Catalunya for their support on this research. This study has been funded by Instituto de Salud Carlos III (ISCIII) through the project “PI21/01019” and co-funded by the European Union. Part of the data acquisition has been supported by PREdICT, sponsored by AstraZeneca. This study has been co-funded by the European Regional Development Fund/European Social Fund ‘A way to make Europe’ (to R.P.L.), and by the Comprehensive Program of Cancer Immunotherapy and Immunology (CAIMI), funded by the Banco Bilbao Vizcaya Argentaria Foundation Foundation (FBBVA, grant 89/2017). R.P.L is supported by the “la Caixa” Foundation CaixaResearch Advanced Oncology Research Program, the Prostate Cancer Foundation (18YOUN19), a CRIS Foundation Talent Award (TALENT19-05), the FERO Foundation through the XVIII Fero Fellowship for Oncological Research, the Instituto de Salud Carlos III-Investigación en Salud (PI18/01395 and PI21/01019), the Asociación Española Contra el Cancer (AECC) (PRYCO211023SERR) and the Generalitat de Catalunya Agency for Management of University and Research Grants of Catalonia (AGAUR) (2023PROD00178). The project that gave rise to these results received the support of a fellowship from “la Caixa” Foundation (ID 100010434). The fellowship code is “LCF/BQ/PR22/11920010” (funding F.G and A.V.). This research has received support from the Beatriu de Pinós Postdoctoral Program from the Secretariat of Universities and Research of the Department of Business and Knowledge of the Government of Catalonia, and the support from the Marie Sklodowska-Curie COFUND program (BP3, contract number 801370; reference 2019 BP 00182) of the H2020 program (to K.B.). M.P. is supported by the UKRI Future Leaders Fellowship MR/T020296/2. A.G. is supported by a Severo Ochoa PhD fellowship (PRE2022-102586). C.M. is funded by the Asociación Española Contra el Cancer (AECC) (PRYCO211023SERR). The authors are thankful to the Vall d’Hebron Radiology department and to the ASCIRES CETIR clinical team for their assistance with MRI acquisitions, and to the GE/Siemens clinical scientists for their support with diffusion sequence characterisation.

## Data and code availability statement

The MC simulation framework upon which *Histo-μSim* has been developed is freely available in GitHub at the permanent address: https://github.com/radiomicsgroup/dMRIMC. The virtual tissue environments reconstructed from histology and the mouse MRI and histology data have been released publicly. The final, permanent addresses of the data bases can be found on the GitHub repository (link: https://github.com/radiomicsgroup/dMRIMC). The data in mouse tissue and in cancer patients used for all MRI-histology correlation analyses are included as tables in this manuscript. Routines for simulation-informed fitting are freely available as part of *BodyMRITools* at the permanent address: https://github.com/fragrussu/bodymritools (script *mri2micro_dictml*.*py*).

## 6 Supplementary material

**Figure S1:**
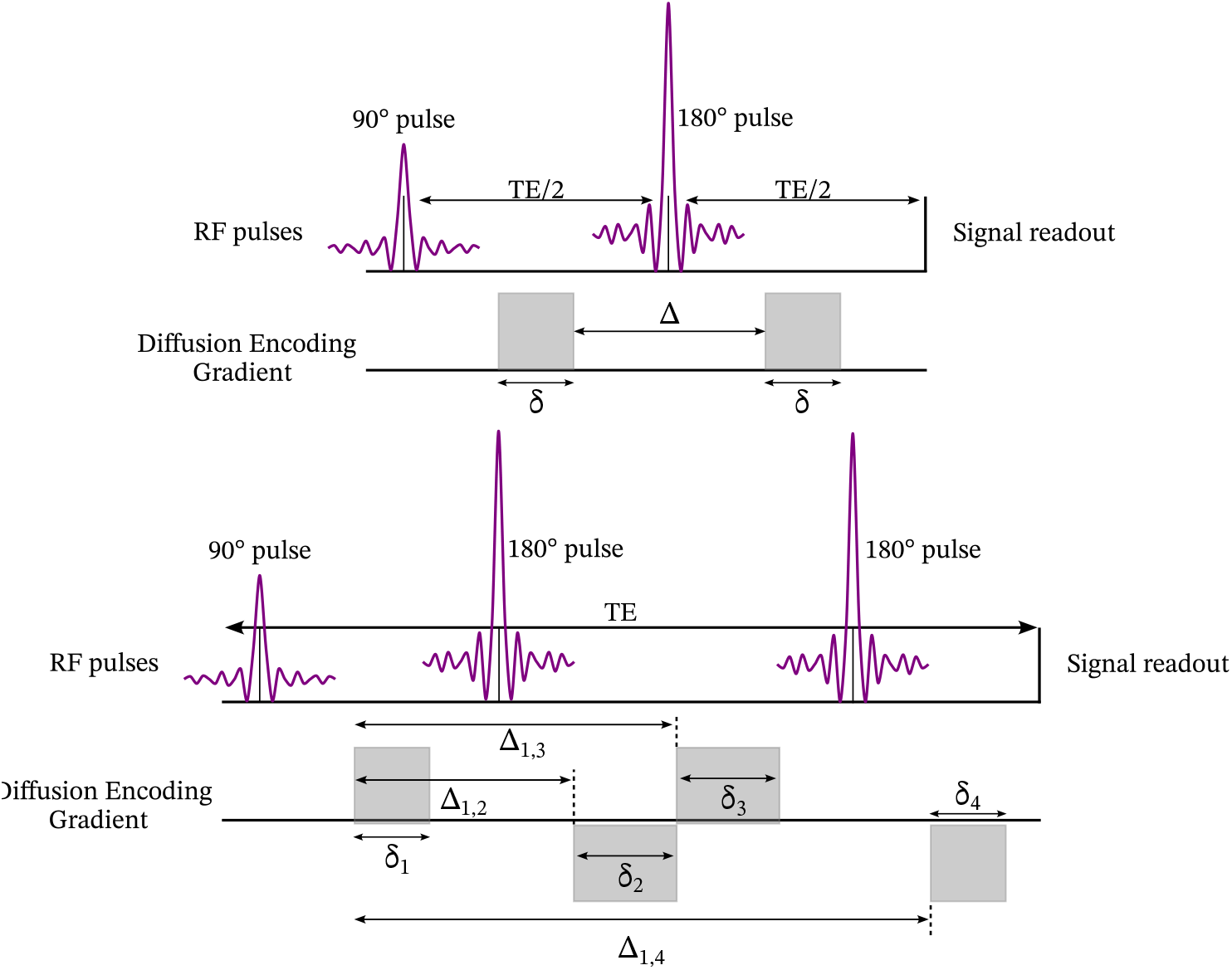
Illustration of the pulsed-gradient spin echo (PGSE, on top) and diffusion-weighted (DW) twice-refocussed spin echo (TRSE, on the bottom) sequences used in this study for both simulations and *ex vivo*/*in vivo* imaging.

**Table S1:** Comparison between histological metrics obtained from manual cell segmentation against automatic segmentation in QuPath (https://qupath.github.io). The table reports the Pearson’s correlation coefficient *r* and the Bias Index (BI) over the 18 mouse data ROIs. BI is defined as the relative percentage difference between QuPath-based values, with respect to manually-derived figures.

| | $f_{in}$ | $vCS_{sph}$ | $vCS_{cyl}$ | mCS | varCS | skewCS |
| --- | --- | --- | --- | --- | --- | --- |
| Pearson's correlation coeff. $r$ | 0.64 | 0.61 | 0.61 | 0.59 | 0.57 | -0.04 |
| Bias Index (BI) (reference: manual) | +5% | +20% | +17% | +11% | +50% | +11% |

**Figure S2:**
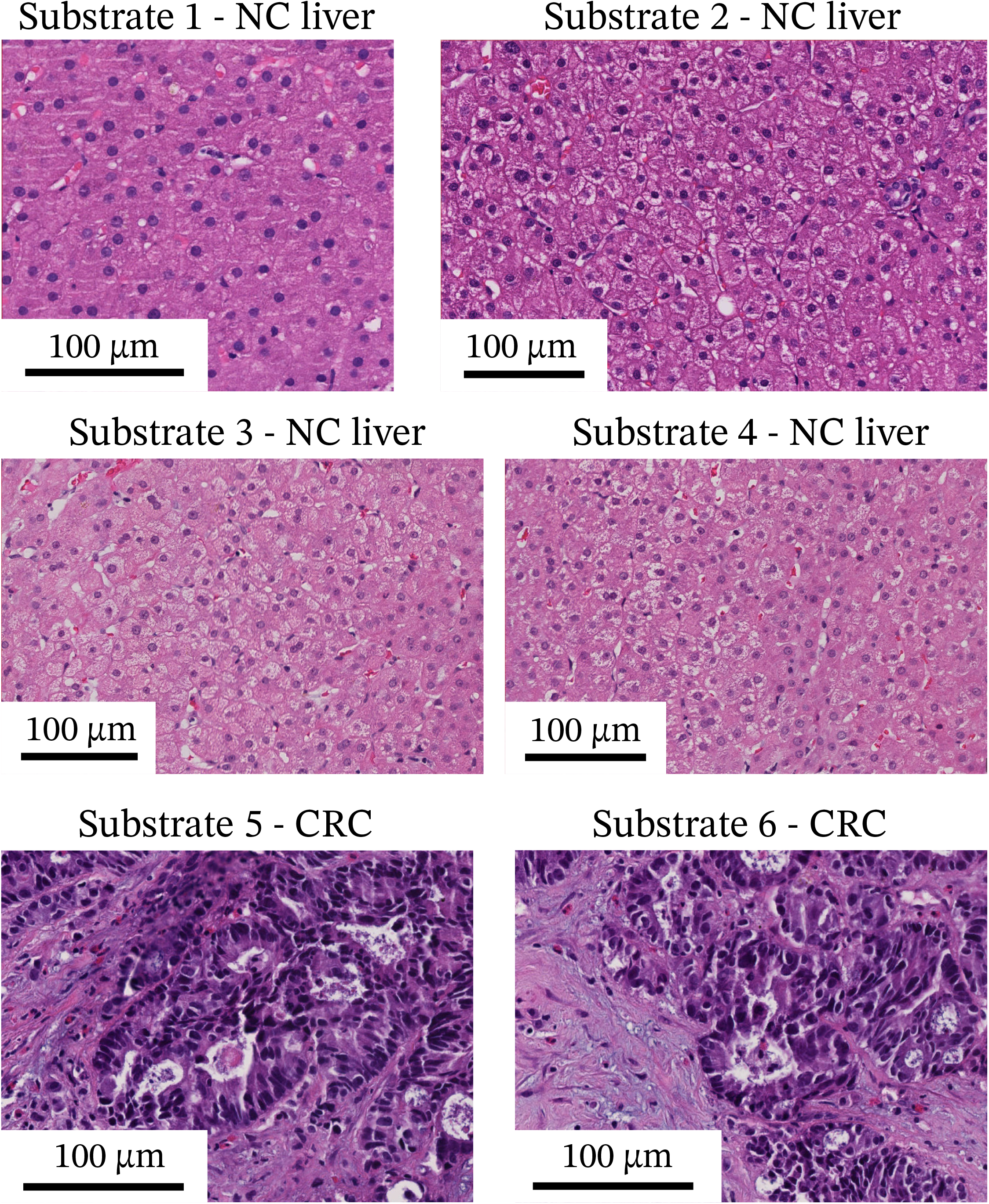
Details of the histological patches from HE-stained biopsies that were manually segmented to reconstruct substrates for Monte Carlo diffusion simulations (substrates 1 to 6). CRC = colorectal cancer; NC = non-cancerous.

**Figure S3:**
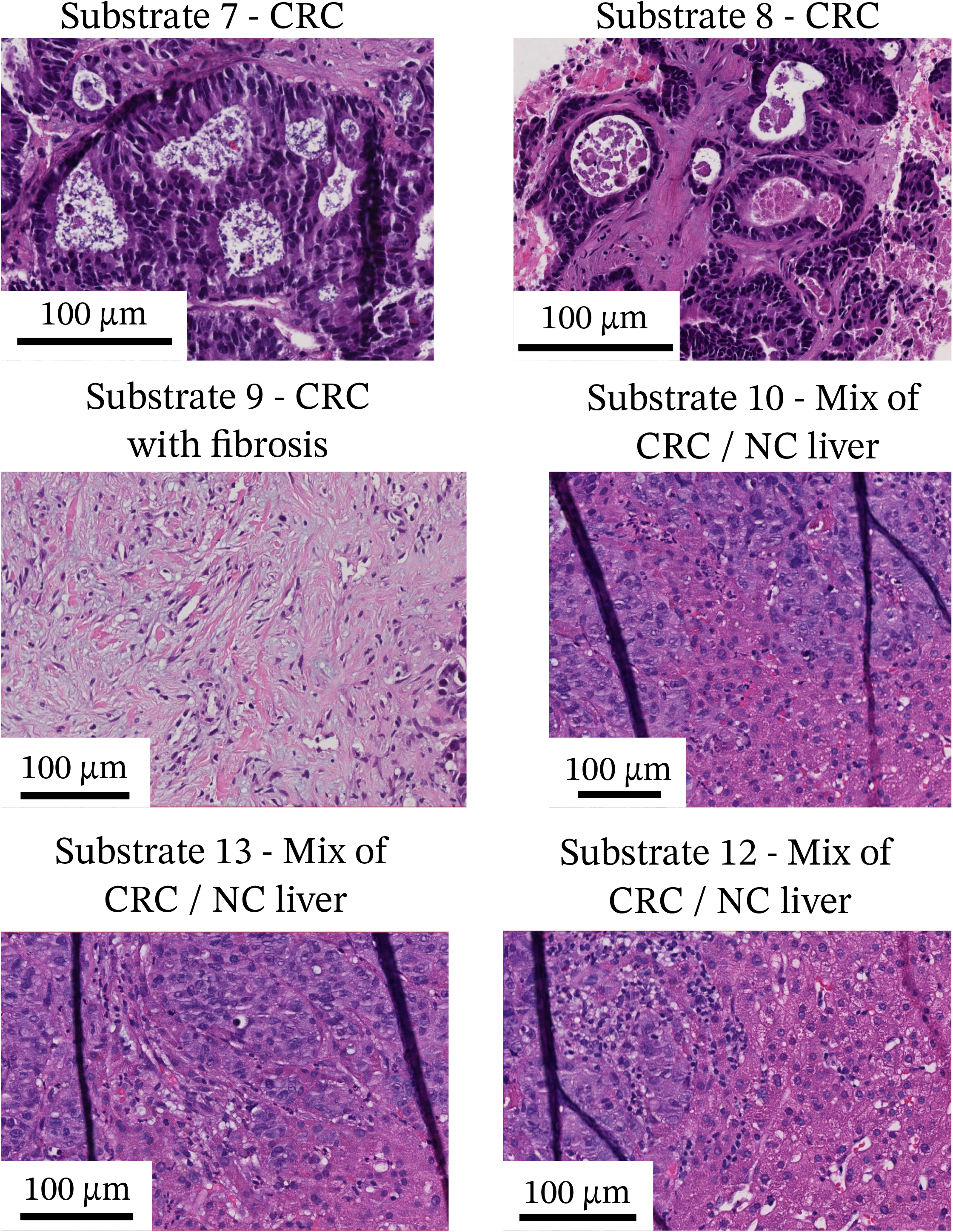
Details of the histological patches from HE-stained biopsies that were manually segmented to reconstruct substrates for Monte Carlo diffusion simulations (substrates 9 to 12). CRC = colorectal cancer; NC = non-cancerous.

**Figure S4:**
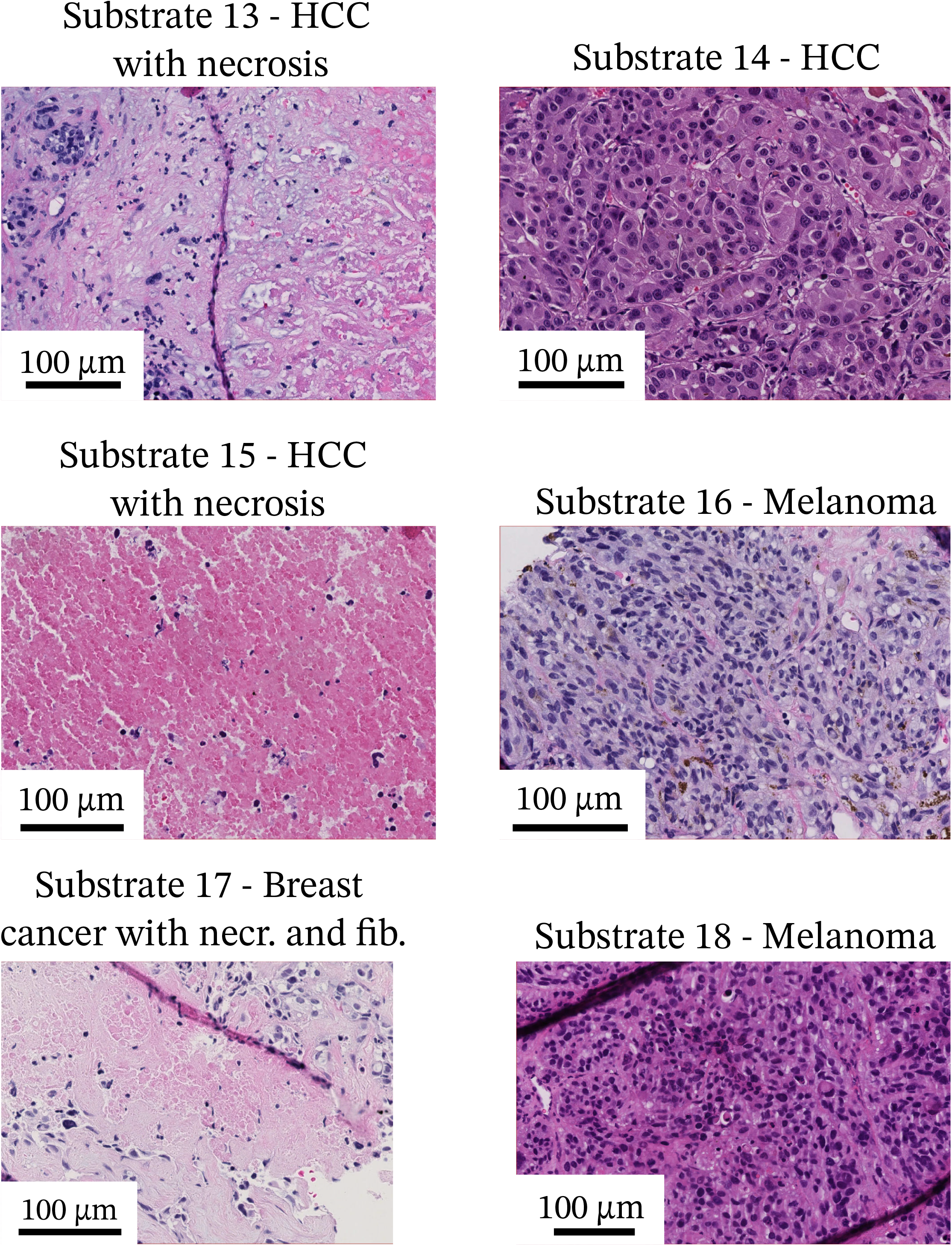
Details of the histological patches from HE-stained biopsies that were manually segmented to reconstruct substrates for Monte Carlo diffusion simulations (substrates 13 to 18). HCC = hepatocellular carcinoma.

**Figure S5:**
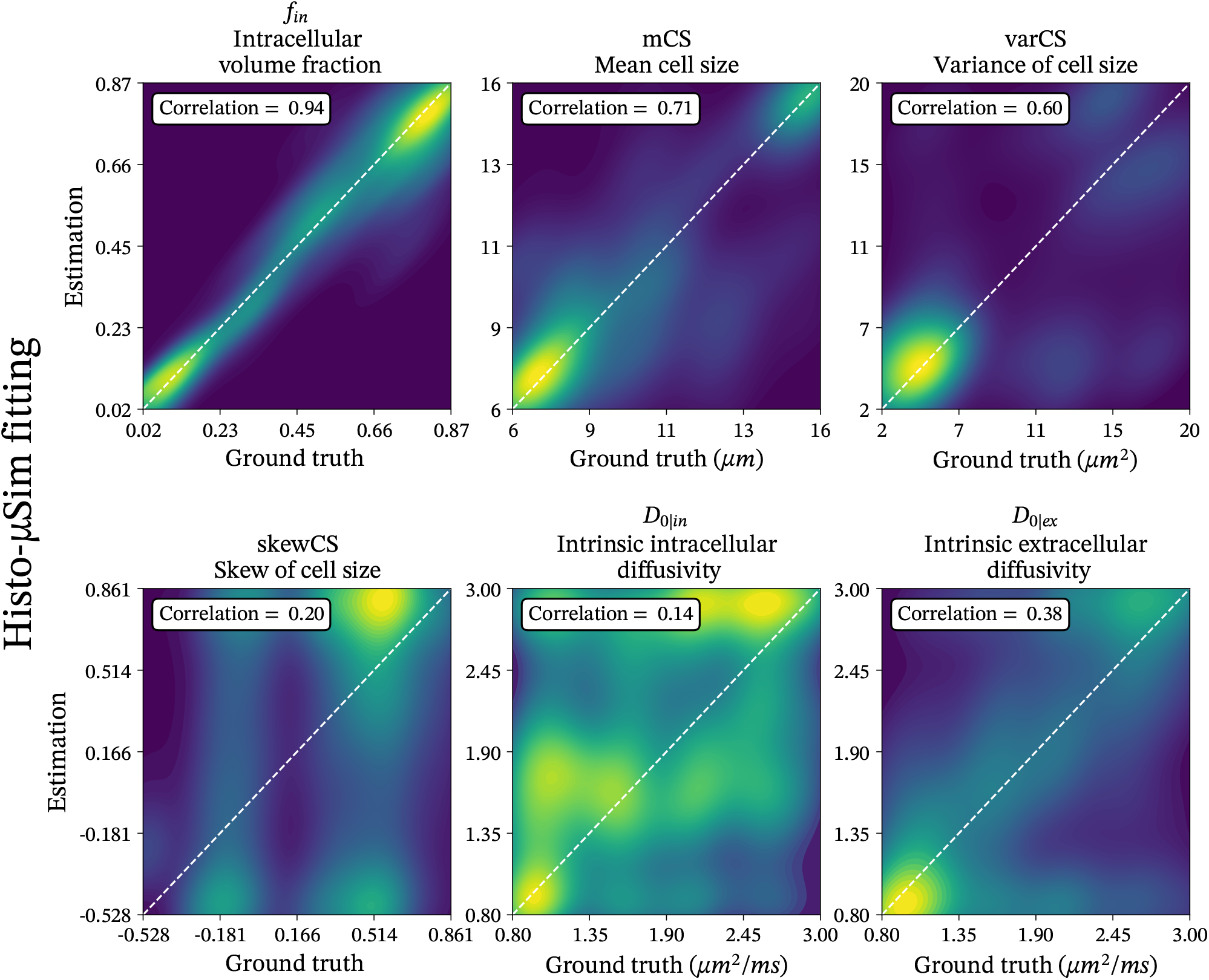
Scatter density plots between ground truth and estimated tissue parameters for MC-informed parameter estimation (forward model 2) and dMRI protocol TRSE. Each plot corresponds to a metric. From the top left corner, in clock-wise order: IC fraction *f*_*in*_, mean CS index, variance of CS varCS, intrinsic EC diffusivity *D*_0|*ex*_, intrinsic IC diffusivity *D*_0|*in*_, skewness of CS distribution skewCS. The plots also include the identity line for reference, and the Pearson’s correlation coefficient between ground truth and estimated parameter values. Results are shown for dMRI protocol *TRSE*.

**Figure S6:**
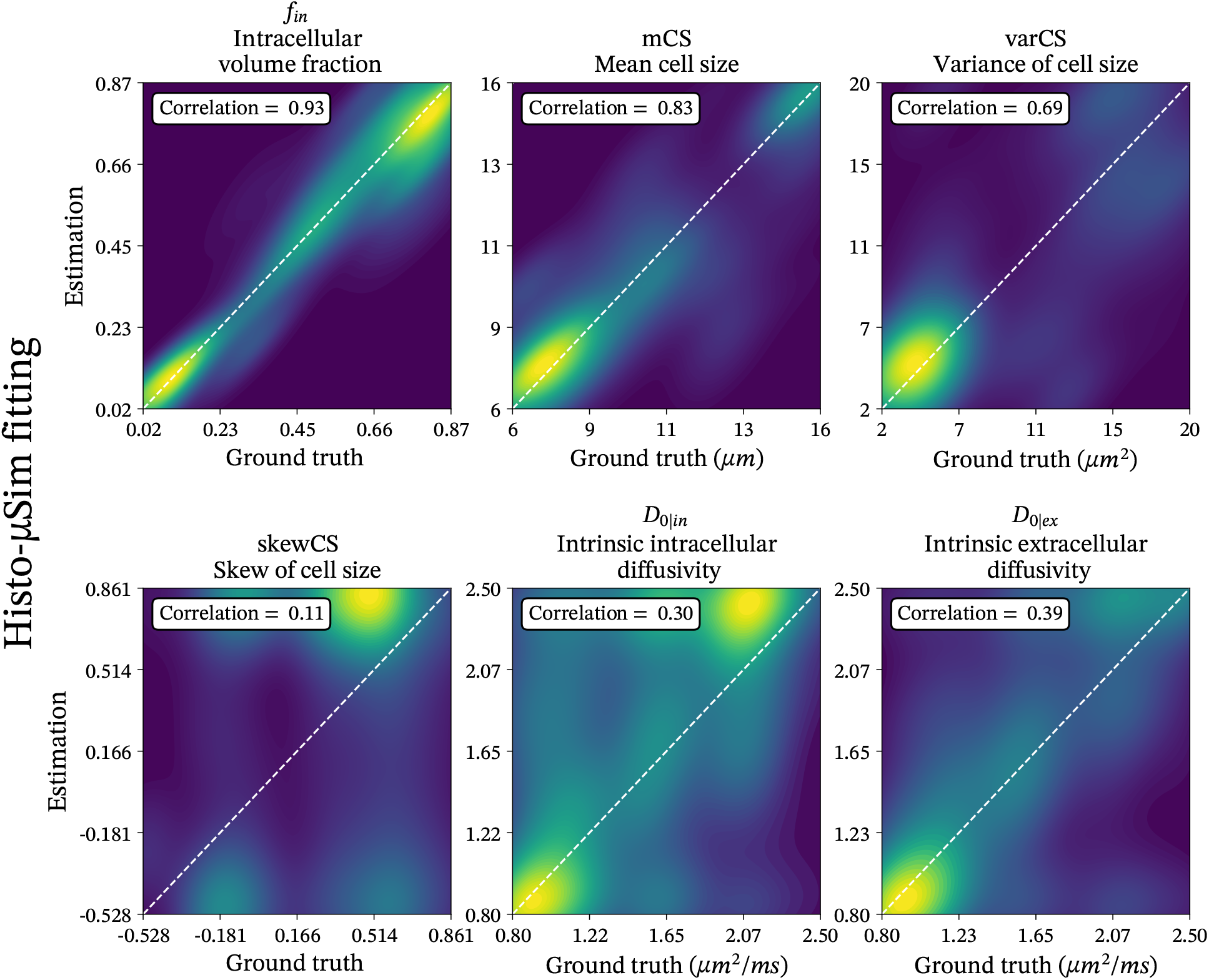
Scatter density plots between ground truth and estimated tissue parameters for *Histo-μSim* MC-informed parameter estimation (forward model 2) and dMRI protocol PGSE-ex. Each plot corresponds to a metric. From the top left corner, in clock-wise order: IC fraction *f*_*in*_, mean CS index, variance of CS varCS, intrinsic EC diffusivity *D*_0|*ex*_, intrinsic IC diffusivity *D*_0|*in*_, skewness of CS distribution skewCS. The plots also include the identity line for reference, and the Pearson’s correlation coefficient between ground truth and estimated parameter values. Results are shown for dMRI protocol *PGSE-ex*.

**Figure S7:**
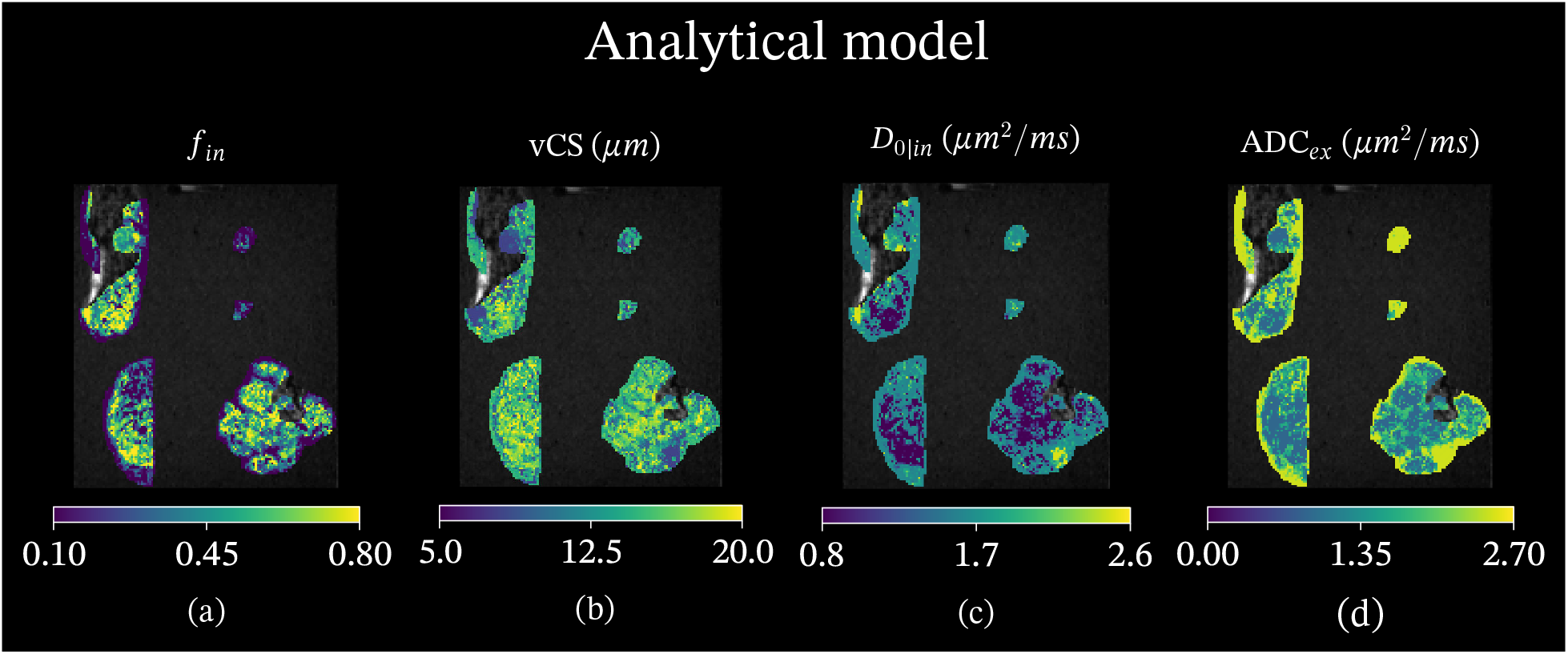
Parametric maps from fitting of the analytical signal model as obtained on the mouse breast specimens scanned *ex vivo* on a 9.4T system. From left to right: IC fraction *f*_*in*_ (a); volume-weighted CS index vCS (b); intrinsic IC diffusivity *D*_0|*in*_ (c); EC apparent diffusion coefficient *ADC*_*ex*_ (f). For each metric, we show results on the four breast specimens. Moving clock-wise: week 9 MMTV-PyM breast tumour (top left), non-cancerous breast (top right), week 11 MMTV-PyM breast tumour (bottom right), week 14 MMTV-PyM breast tumour (bottom left).

**Figure S8:**
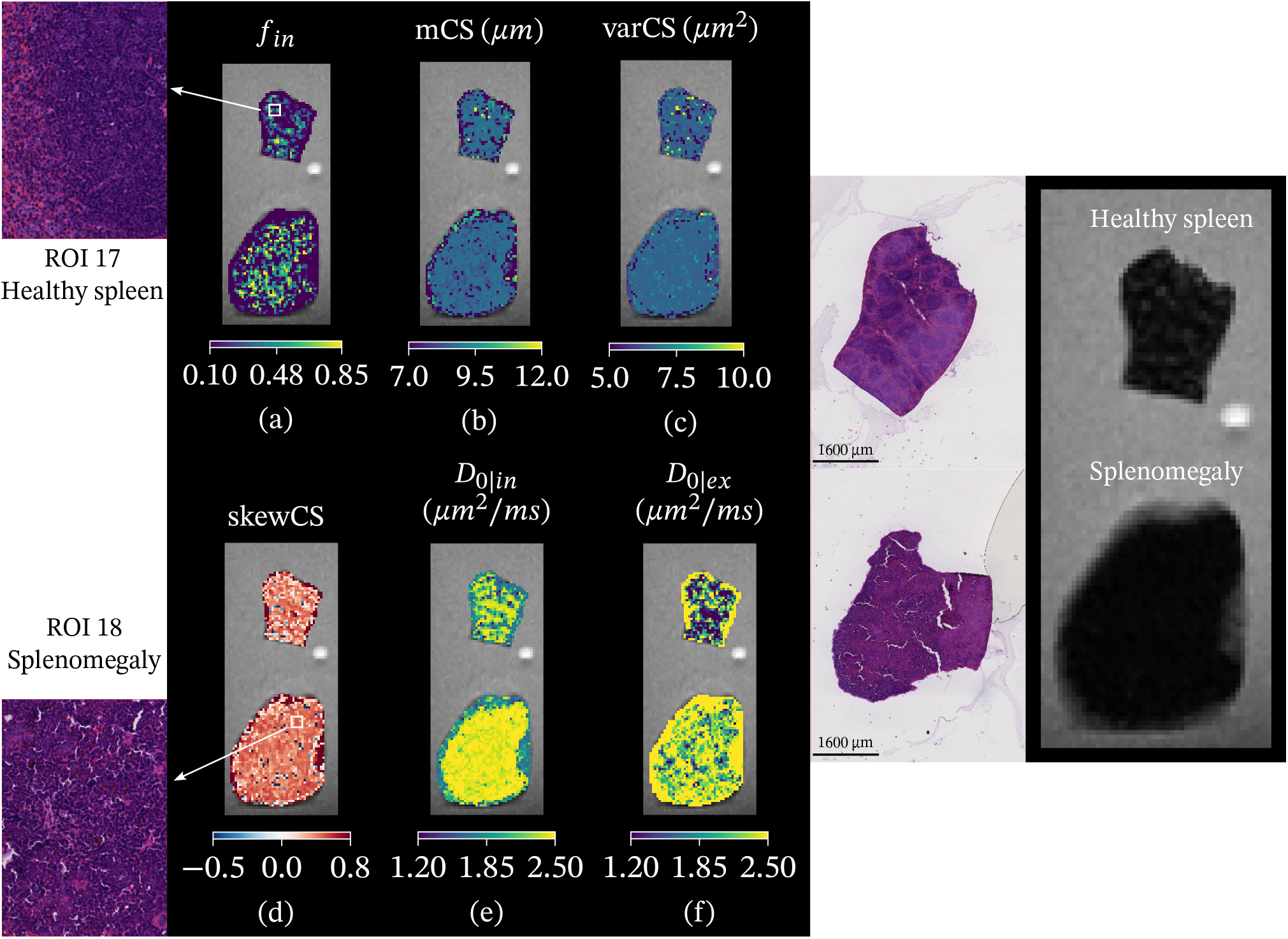
*Histo-μSim* maps (forward model 2) and histological results on two fixed mouse spleens scanned *ex vivo* on a 9.4T system. Right: *b* = 0 image, co-localised HE-stained section, and high-resolution histological patches. Left: parametric maps from forward model 2. First row: IC fraction *f*_*in*_ (a); mean CS index mCS (b); variance of CS varCS (c). Second row: skewness of the CS distribution skewCS (d); intrinsic IC diffusivity *D*_0|*in*_ (e); intrinsic EC diffusivity *D*_0|*ex*_ (f). For each metric, we show results on both samples: Normal spleen (top), splenomegaly (bottom).

**Figure S9:**
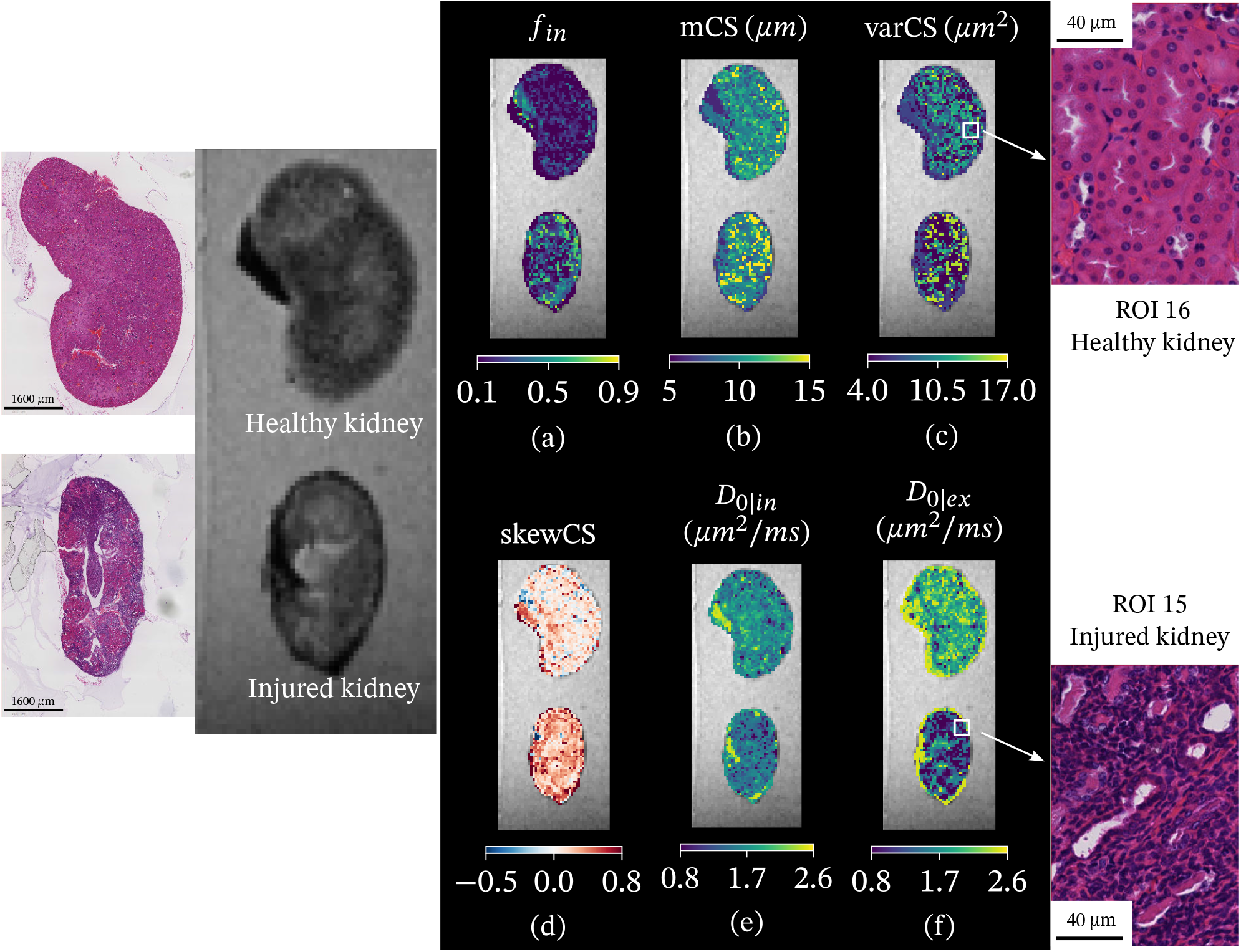
*Histo-μSim* maps (forward model 2) and histological results on two fixed mouse kidneys scanned *ex vivo* on a 9.4T system. Left: *b* = 0 image, co-localised HE-stained section, and high-resolution histological patches. Right: parametric maps from forward model 2. First row: IC fraction *f*_*in*_ (a); mean CS index mCS (b); variance of CS varCS (c). Second row: skewness of the CS distribution skewCS (d); intrinsic IC diffusivity *D*_0|*in*_ (e); intrinsic EC diffusivity *D*_0|*ex*_ (f). We show again results on both samples: Normal kidney (top), folic-acid induced kidney injury (bottom).

**Figure S10:**
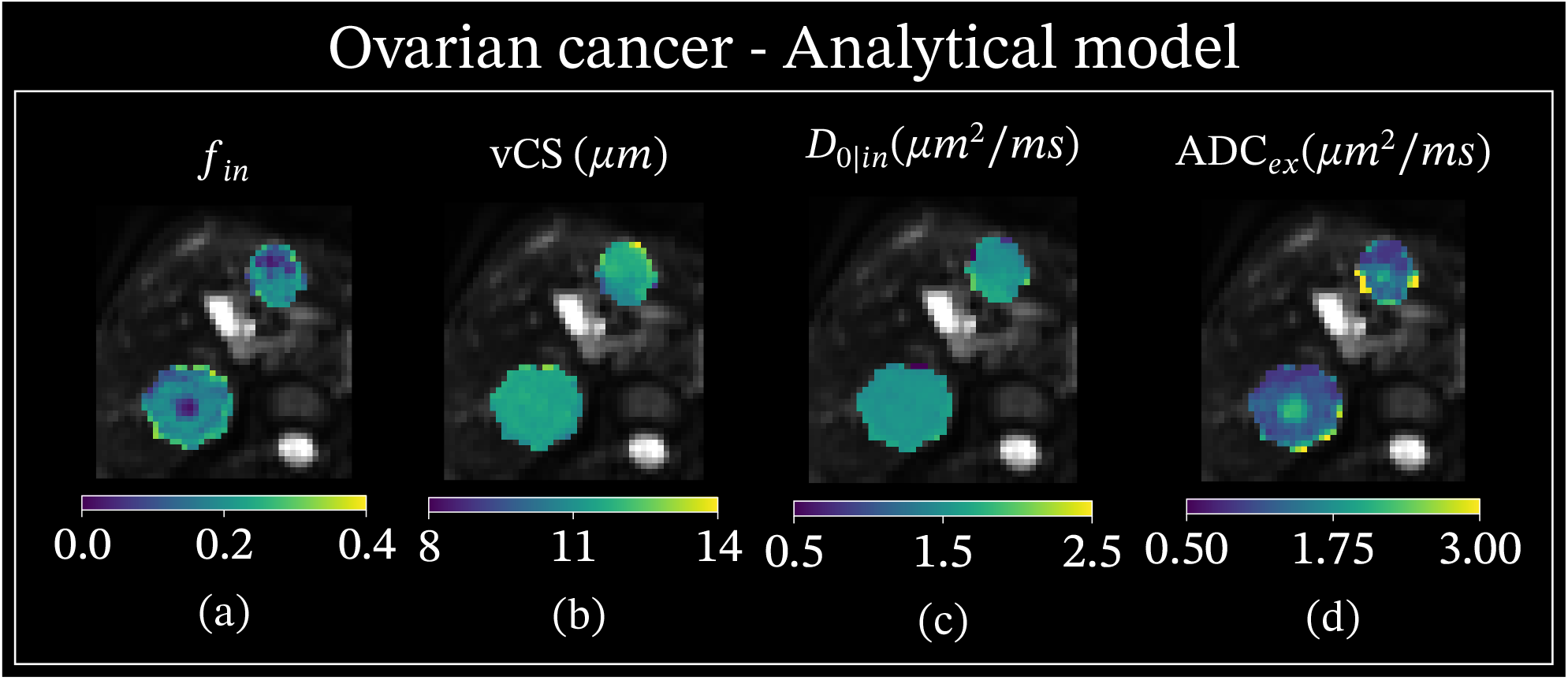
Parametric maps from the analytical signal model obtained on a representative patient scanned on the 3T system (ovarian cancer liver metastases; same case shown for *Histo-μSim* in Fig. 10). From left to right: *f*_*in*_ (a), vCS (b), *D*_0|*in*_ (c), *ADC*_*ex*_ (d).

**Figure S11:**
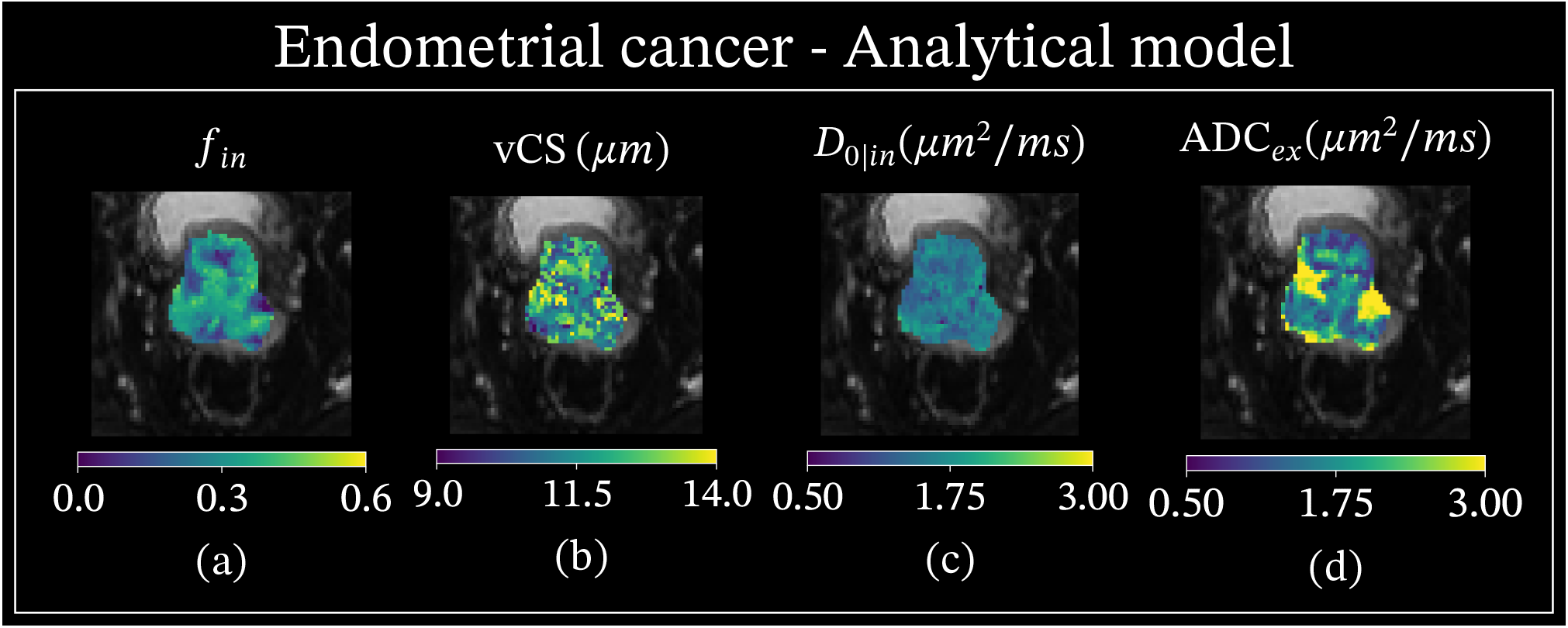
Parametric maps from the analytical signal model obtained on a representative patient scanned on the 1.5T system (endometrial cancer; same case shown for *Histo-μSim* in Fig. 10). From left to right: *f*_*in*_ (a), vCS (b), *D*_0|*in*_ (c), *ADC*_*ex*_ (d).

